# A novel approach to concentrate human and animal viruses from wastewater using receptors-conjugated magnetic beads

**DOI:** 10.1101/2021.12.07.21267392

**Authors:** Chamteut Oh, Kyukyoung Kim, Elbashir Araud, Leyi Wang, Joanna L. Shisler, Thanh H. Nguyen

## Abstract

Viruses are present at low concentrations in wastewater, and therefore an effective concentration of virus particles is necessary for accurate wastewater-based epidemiology (WBE). We designed a novel approach to concentrate human and animal viruses from wastewater using porcine gastric mucin-conjugated magnetic beads (PGM-MBs). We systematically evaluated the performances of the PGM-MBs method (sensitivity, specificity, and robustness to environmental inhibitors) with six viral species including Tulane virus (a surrogate for human norovirus), rotavirus, adenovirus, porcine coronavirus (transmissible gastroenteritis virus or TGEV), and two human coronaviruses (NL63 and SARS-CoV-2) in influent wastewater and raw sewage samples. We determined the multiplication factor (the ratio of genome concentration of the concentrated over that of the initial solution) for the PGM-MBs method, which ranged from 1.3 to 64.0 depending on the viral species. Because the recovery efficiency became significantly higher when calculated based on virus titers than genome concentration, the PGM-MBs method could be an appropriate tool for assessing the risk due to wastewater contaminated with infectious enteric viruses. PCR inhibitors were not concentrated by PGM-MBs, suggesting this tool will be successful for use with environmental samples. The PGM-MBs method is cost-effective (0.43 USD/sample) and fast turnaround (3 hours from virus concentration to genome quantification), and thus this method can be implemented for high throughput facilities. Based on good performance, intrinsic characteristics of targeting the infectious virus, robustness to wastewater, and adaptability to high throughput systems, we are confident that the PGM-MBs method can be applied for successful WBE and ultimately provides valuable public health information.

**Graphical abstract:** 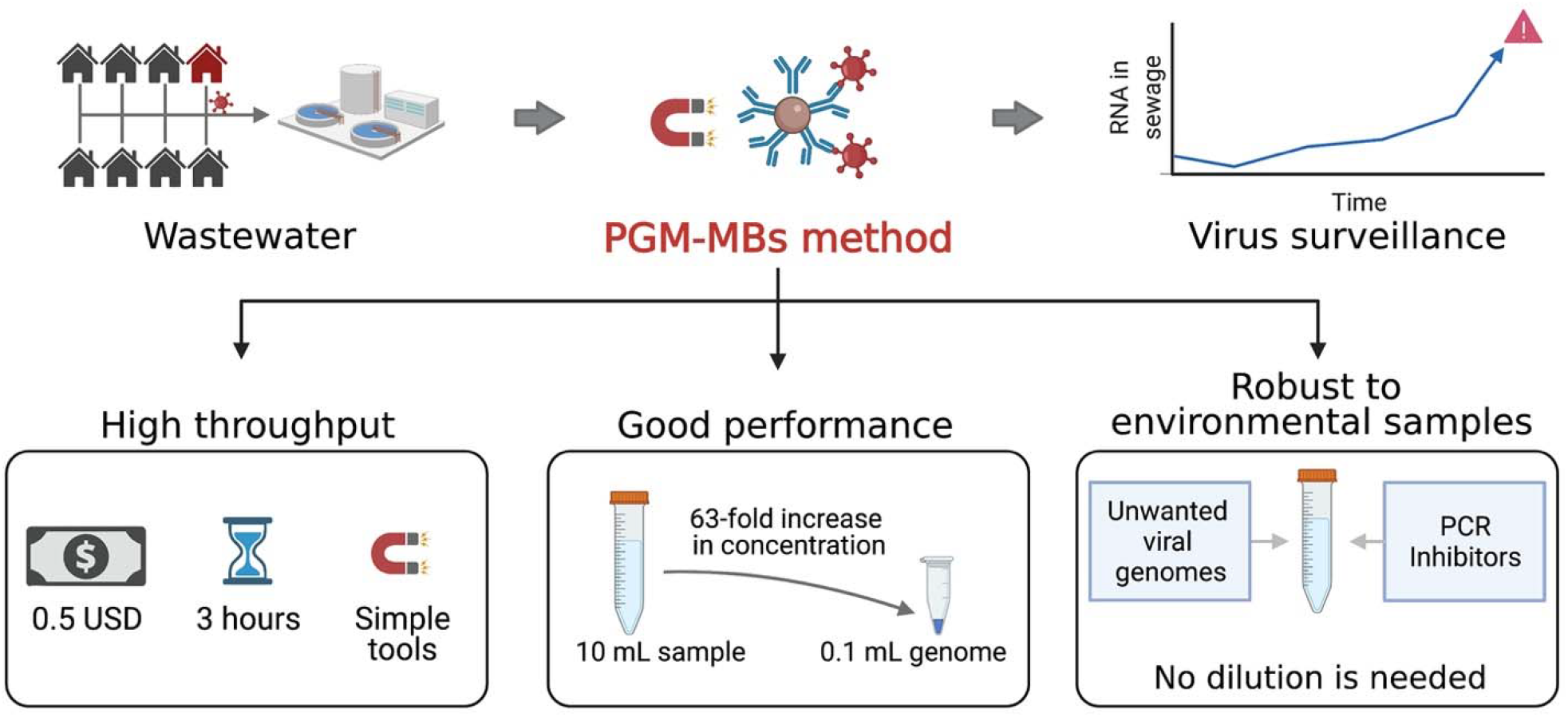

## 1. Introduction

Enteric viruses are the leading cause of gastroenteritis worldwide. Many enteric viruses, such as rotavirus, norovirus, and adenoviruses, are shed in fecal material (Kang, 2017; Kapikian, 1996). Other airborne viruses, like SARS-CoV-2, are also shed in the human stool (Hu et al., 2020). Thus, wastewater-based epidemiology (WBE) can be a powerful tool to detect viral transmission in communities even before the appearance of clinical cases (Ahmed et al., 2021; Harris-Lovett et al., 2021; Hart and Halden, 2020; Nemudryi et al., 2020; Panchal et al., 2021; Sherchan et al., 2021). However, virus monitoring faces a challenge that the viral concentration in wastewater may be below the detection limit of analysis instruments such as qPCR system, especially when transmission has just started in communities. Although viruses such as noroviruses, adenoviruses, and enteroviruses are excreted at high concentrations (up to 10^11^ viruses/g-feces), the viruses are diluted by different sources of water (Haramoto et al., 2018). For example, Albinana-Gemenez et al. (2006) detected 10^6^-10^7^ gene copies (gc)/L of adenoviruses and polyomaviruses in sewage, but the virus concentrations decreased to 10^1^ to 10^2^ gc/L for river water. Since the lower limit of qPCR detection is about 10^0^ gc/μL (or 10^6^ gc/L), viruses from wastewater samples must be concentrated into smaller volumes. There are several strategies available to concentrate viruses from wastewater include ultracentrifugation, ultrafiltration, adsorption/extraction, skimmed-milk flocculation, polyethylene glycol (PEG) precipitation, and sludge extraction methods (Ahmed et al., 2020; Jafferali et al., 2021; Wolfe et al., 2021). However, those conventional virus concentration methods are expensive, time-consuming, and resource-demanding processes (Gibas et al., 2021; LaTurner et al., 2021; Polo et al., 2020), so this process often becomes a bottleneck in monitoring viruses in wastewater. Therefore, there is an urgent need for a new virus concentration method that is simple and fast.

In the recent past, we and others have coated magnetic beads (MB) with porcine gastric mucin (PGM) to study virus binding properties (Afolayan et al., 2016; Araud et al., 2018; Fuzawa et al., 2019; Oh et al., 2020; Walker et al., 2019). The PGM is a biological substrate that includes molecules (e.g., glycans) that act as virus receptors. Thus, we hypothesize that these PGM-MBs can selectively concentrate viruses from wastewater. An additional benefit of PGM-MBs is that they can capture infectious virus particles instead of viral genomes, giving a more accurate quantification of risk for WBE.

The objective of this research was to design and optimize this receptor-based virus concentration method using porcine gastric mucin conjugated-magnetic beads (PGM-MBs) and evaluate the efficacy of PGM-MBs compared to its conventional counterparts. **Fig. S1** shows five steps of the PGM-MBs method from wastewater sample pretreatment, virus attachment, virus concentration, genome extraction, to genome quantification step. We first optimized the method using influent wastewater spiked with five virus strains (Tulane virus (TV), rotavirus (RV), adenovirus (AdV), human coronavirus (NL63), and porcine coronavirus (transmissible gastroenteritis virus; TGEV). We selected these viruses based on their significance for public health and economy (Binder et al., 2017; Fielding, 2011; Katayama et al., 2002; La Rosa et al., 2020a; Lodder and De Roda Husman, 2005; Waruhiu et al., 2017). We systematically evaluated the performances of the PGM-MBs method (sensitivity, specificity, and robustness to environmental inhibitors) with 20 sewage samples spiked with these enteric viruses. In addition, we used PGM-MBs in local sewage samples to concentrate SARS-CoV-2 and compared its performances to electronegative filtration methods, one of the most widely used virus concentration methods. The findings suggest that PGM-MBs can be simple, affordable, but still accurate tools for WBE.

## 2. Materials and Methods

### 2.1. PGM-MBs virus concentration method

The porcine gastric mucin conjugated magnetic beads (PGM-MBs) were produced as described in our previous studies (Araud et al., 2018; Fuzawa et al., 2019; Oh et al., 2020), and the detailed procedure is elaborated in **Text S1**. The PGM-MBs is suspension that includes 1-4 µm diameter beads at a concentration of 2×10^6^ beads/µL. The pretreatment of wastewater samples, necessary to remove solid particles that can otherwise interference with subsequent analysis, was achieved by either gravitational settling for 2 hours or filtration through 0.22 μm membrane filters (S2GPT02RE, Millipore Sigma, USA). The virus attachment to the PGM-MBs was optimized by changing the MgCl_2_ concentration between 0 to 100 mM. Specifically, the PGM-MBs and MgCl_2_ were added to a wastewater sample at a 1:1000 volume ratio and 50 mM, respectively. For example, we put 10 µL of the PGM-MBs into 10 mL of the samples for the optimization experiments and 50 µL PGM-MBs to 50 mL sewage for SARS-CoV-2 surveillance. The mixture was shaken for 30 minutes on an orbit shaker at 450 rpm (Fisher Scientific, USA) at room temperature. Then, the virus-containing PGM-MBs were concentrated by a magnet (DH125J-FN, Amazing magnets, USA) placed at the outside surface of the bottom of the tube for about 30 minutes and the clear supernatant was removed. The remaining PGM-MBs and supernatant were collected and transferred to a 1.5 mL low adhesion microcentrifuge tube (1415-2600, USA scientific, USA). The PGM-MBs were washed three times by sequentially resuspending beads in 1 mL PBS, collecting beads by using a magnetic separation rack (S1509S, New Biolabs Lab, USA), and removing supernatants. Finally, the viral genomes were extracted by heat denaturation method and quantified by either qPCR or RT-qPCR depending on viral species, which is elaborated in Section 2.1. Also, Section 3.1 presents the results of the PGM-MBs optimization experiments.

### 2.2. Viral nucleic acid extraction and quantification

Commercial nucleic acid extraction kits are expensive, time-consuming and could be in short supply (Satyanarayana, 2020). Therefore, we established a heat denaturation method compatible with the PGM-MBs method to reduce resources (cost, time, and labor). The heat denaturation method started with resuspending a pellet of virus-containing PGM-MBs with 100 μL proteinase K (P8107S, New England Biolabs, USA) that was diluted 100-fold by molecular biology grade water (Millipore Sigma) for denaturation of the viral capsid. The mixture was then transferred to a 200 μL-size PCR tube (14-222-262, Fisher Scientific, USA) and incubated at 95□ for 10 minutes using a thermal cycler (MyCycler™, Bio-Rad), followed by quick cooling on ice. While being heated, the viral genomes were released from the capsid, and the proteinase K was inactivated. The PGM-MBs were pelletized by a magnet (S1509S, New Biolabs Lab, USA) for one minute, and the genomes were taken from the supernatant near the solution surface for further analysis. The concentrations of viral genomes were quantified by SYBR-based RT-qPCR for TV, RV, NL63, and TGEV; Taqman-based RT-qPCR for SARS-CoV-2;, or SYBR-based qPCR for AdV. We used iTaq universal SYBR green reaction mix (Bio-Rad Laboratories, USA) for TV, RV, NL63, and TGEV analysis, PowerUp SYBR(tm) Green Master Mix (Applied Biosystems, CA, USA) for AdV quantification, and Taqman Fast Virus 1-step Master Mix (4444432, Applied Biosystems, USA) for SARS-CoV-2. Viral genomes were quantified using a qPCR system (QuantStudio 3, Thermo Fisher Scientific, USA). **Table S1** summarized these RT-qPCR methods following the MIQE guidelines (Bustin et al., 2009). **Text S2** provides detailed information about qPCR analysis, including thermal cycles, qPCR cocktail compositions, and primers.

### 2.3. Preparing viruses for experiments with virus-spiked wastewater

We determined the performance of the PGM-MBs method using influent wastewater spiked with viruses propagated *in vitro*. These viruses are Tulane virus (TV, *Caliciviridae*, a viral surrogate for human norovirus) (Yu et al., 2013), rotavirus OSU strain (RV, *Reoviridae*, ATCC VR-892), adenovirus type 2 (AdV, *Adenoviridae*, ATCC VR-846), human coronavirus (NL63, *Coronaviridae*), and porcine coronavirus (TGEV, *Coronaviridae*). We obtained TV from the Cincinnati Children’s Hospital Medical Center. We obtained NL63 strain from BEI sources (NR-470). TGEV is a porcine enteric virus, and is in the same family as NL63 and SARS-CoV-2 (*Coronaviridae*). We received the TGEV from the Veterinary Diagnostic Laboratory at the University of Illinois at Urbana-Champaign. Rotavirus strain OSU strain (RV, *Reoviridae*, ATCC VR-892) and adenovirus type 2 (AdV, *Adenoviridae*, ATCC VR-846) were obtained from the ATCC. Detailed information about propagation methods including culture media and growing conditions is elaborated in **Text S3**. Plaque assays were used to determine titers of TV, RV, AdV, and TGEV. TCID50 assays were used to quantify NL63 titers. MA104 (ATCC, CRL-2378.1) was used as a host cell line for TV and RV plaque assay. A549 (ATCC; CCL-185), MK2 cells (ATCC, CCL-7), and ST cells (CRL-1746, ATCC) were used to determine virus titers of AdV, NL63, and TGEV, respectively. Detailed information on virus titer determination is summarized in **Text S4**.

### 2.4. Collection of influent wastewater and local sewage samples for method optimization and evaluation

We collected environmental samples from 10 locations from January to July 2021 (20 samples in total) to cover a variety of wastewate. We collected a 24-hour composite of influent wastewater on February 3rd, 2021, from the Urbana Ȧ Champaign Sanitary District, IL, USA, a district that serves about 100,000 local residents. Because the bacteria present in the wastewater interfere with the plaque assays, a portion of the influent wastewater was filtered through a 0.22 μm polyethersulfone membrane filter (S2GPT02RE, Millipore Sigma, USA), and the filtered influent wastewater was used for sensitivity and specificity experiments (Section 3.1, 3.2, 3.3, and 3.6). The filtrate was stored at -80□ until used. The filtrate was thawed and stored at 4□ for less than two weeks, a time period where we conducted all experiments in this study.

Three-day composite sewage samples were collected from January to July 2020 from 10 different neighborhood-level sewersheds across Champaign County, IL, USA (**Table S3**). These sewage samples were stored at 4□ for less than two weeks before being used to test the impact of the filtration process on performances of the PGM-MBs method (Section 3.4) and the tolerance of the PGM-MBs method to PCR inhibitors (Section 3.5). In addition, the SARS-CoV-2 concentration in these sewages were measured for a smaller aliquot of these samples kept at -80□ for less than a week and analyzed as soon as they were defrosted (Section 3.7). Information including sampling locations and dates was summarized in **Table S3**.

### 2.5. Recovery effliencies of the PGM-MBs method

In an experiment conducted to evaluate performances of the PGM-MBs method with different viral genome concentrations, we spiked 100 μL of either TV, RV, AdV, NL63 or TGEV into 10mL of filtered influent wastewater (**Table S3**). The initial genome copies spiked to wastewater were about 10^7^ gc for TV and NL63 and about 10^8^ gc for RV, AdV, and TGEV. The wastewater was serially diluted in 10-fold increments (four serial dilutions for TV and NL63 and five serial dilutions for RV, AdV, and TGEV) and the three biological replicates were prepared. The results from this experiment can be found in Section 3.2.

We conducted another experiment to compare the recovery efficiencies (RE) of the PGM-MBs method by examining viral genome concentration or virus titer. For these experiments, 10 μL of virus (about 10^4^ PFU of TV, RV, AdV, and TGEV, and 10^3^ TCID50 of NL63) and 10 μL of MgCl_2_ (final concentrations of 50 mM) was added to 1 mL of six different filtered sewage samples (**Table S3**). Viral genome concentrations and infectious virus titers in the initial sewage samples were quantified. After that, we put 10 μL of PGM-MBs to these sewage samples. Then, we took the supernatant and quantified its genome concentrations and virus titers. We assumed that the difference in viral genomes and virus titers between the initial sewage and the supernatant is the recovered genomes and virus titers by the PGM-MBs method (Eq. 1). Section 3.3 presents the results from this experiment.

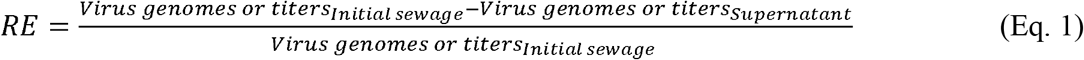

### 2.6. Tolerance for PCR inhibitors

We conducted a set of experiments to understand the impact of solid particles on performances (RE and tolerance to PCR inhibitors) of the PGM-MBs method. We chose 6 sewage samples collected from different sampling locations or times. These sewage samples were left in a refrigerator at 4□ for two hours without disturbance to mimic sedimentation. The liquid near the surface of these samples was used for two experiments. In the first experiment, this liquid was used as it is. In the second experiment, this liquid was filtered by 0.22 μm pore size filters. Each of five viral species (about 10^6^ gc) was added to 10 mL of either the filtered or the unfiltered samples before these samples were subjected to the PGM-MBs. The concentrations of viral genomes in samples before and after the PGM-MBs method were quantified and the recovery efficiencies were calculated. In addition, we tested the impact of solid particles as PCR inhibitors. In these experiments, we used the same pairs of unfiltered and filtered samples described above. Then, we applied the PGM-MBs method to the sewage samples, to which we did not spike any viruses. After we obtained the final extracts from the PGM-MBs sample, we spiked 1 μL of TV genomes to 10 μL of the final extracts. Because TV is a Rhesus monkey virus, they are not expected in the sewage samples. The same number of TV genomes was added to PCR inhibitor-free water (molecular biology grade water) as control samples. The results about the impact of solid particles on the performances of the PGM-MBs method are presented in Section 3.4.

We also tested tolerance of the PGM-MBs method to PCR inhibitors in environmental samples. We collected 19 different sewage samples and 1 sample from lagoon (**Table S3**). We also dissolved humic acid (41747, Alfa Aesar, USA), which was extracted by alkaline extraction method from brown coal, to molecular biology grade water at a final concentration of 20 mg C/L. These 21 samples were filtered through 0.22 μm membrane filters before being processed these 21 samples with two different methods: a commercially available genome extraction kit (Viral RNA Mini Kit) and the PGM-MBs method. We subjected 140 μL of the samples to the Viral RNA Mini Kit (QIAGEN, German) and obtained 60 µL of the final solution. To test the PGM-MBs method, we put 10 μL of the PGM-MBs to 10 mL of the samples adjusted to 50 mM of MgCl_2_ and 100 μL of a final solution was obtained. Then, we spiked 1 μL of TV genomes to 10 μL of the two types of final solutions (i.e., genome extraction kit and PGM-MBs method) and molecular biology grade water as PCR inhibitor negative controls. The results on the tolerance of the PGM-MBs to PCR inhibitors are presented in Section 3.5.

### 2.7. Specificity of the PGM-MBs method

We prepared three different solutions: molecular biology grade water, filtered influent wastewater without spiking viruses, and filtered influent wastewater with the four different viral species spiked. For example, for a TV specificity test, the other four virus species (RT, AdV, NL63, and TGEV) were spiked into 10 mL of the filtered influent wastewater to test impact of co-existing viral species on detection of TV genomes. Then, we applied the PGM-MBs method to each of these three solutions and analyzed target virus concentrations in the final extracts from the control wastewater and the four virus-spiked wastewater. All samples were categorized into either positives (i.e., Ct values less than 40) or negatives (undetermined or Ct values higher than 40). The results of the specificity experiment can be found in Section 3.6.

We applied the PGM-MBs method and electronegative membrane filtration method to 50 mL of 7 different sewage samples. The sewage samples were filtrated by 0.22 μm membrane filters and the filtrate was supplemented by the MgCl_2_ at a final concentration of 50 mM. We added about 10^5^ gc of NL63 as a internal control. These samples were incubated at room temperature for 30 minutes before processing them either by the PGM-MBs method or the electronegative membrane filtration method. For the PGM-MBs method, we put 50 μL of the PGM-MBs to the 50 mL of samples. The electronegative membrane filtration started with filtering the 50 mL of sewage samples through 0.45 μm electronegative membranes. After that, the membrane filter was placed into a 5 mL tube (0030119487, Eppendorf, Germany). We put a mixture of lysing buffer (2800 μL) and RNA carrier (28 μL) followed by 1-minute vortex, 15-minute shaking at 450 rpm, and subsequent 1-minute vortex. Section 3.7 presents the results of the comparison between the PGM-MBs method and the electronegative membrane filtration method.

### 2.8. Statistical analysis

Mann-Witney test was conducted to compare differences in recovered viral genomes depending on different MgCl_2_ concentraions (**Fig. 1A**) and extraction method (i.e., heat denaturation method and extraction kit; **Fig. 1B**). Paried sample t-test was performed to compare recovery efficiencies determined either by genome concentration or virus titer (**Fig. 5**) and to compare recovery efficiencies with filtered and unfiltered sewage samples (**Fig. 6**). There were no outliers (i.e., all data for the paired t-test exist within a range from Q1-1.5IQR and Q3+1.5IQR). Also, normalities of the differences between two data sets were checked by Shapiro-Wilk test. Two-sample proportion test was conducted to compare two binomial proportions of Ct values less than 40 from control wastewater and virus-spiked wastewater in **Table 2**. We added notes to **Table 2** showing if sample size (n) times proportion (p) is greater than 5, which is an assumption for the two-sample proportion test. All statistical analysis was conducted by OriginPro 2019b.

**Table 1.** The summary of the virus concentration experiment using the PGM-MBs method with 10 mL of filtered influent wastewaters

| Viral species | RE | CF | MF | LOQ of PGM-PMs method (Log<sub>10</sub> gc/L) |
| --- | --- | --- | --- | --- |
| TV | 0.152±0.092 | 0.01 | 15.2±9.2 | 5.10 |
| RV | 0.123±0.041 | 0.01 | 12.3±4.1 | 5.57 |
| AdV | 0.640±0.166 | 0.01 | 64.0±16.6 | 4.63 |
| **NL63** | 0.103±0.023 | 0.01 | 10.3±2.3 | 5.62 |
| **TGEV** | 0.013±0.005 | 0.01 | 1.3±0.5 | 6.08 |

**Table 2.**
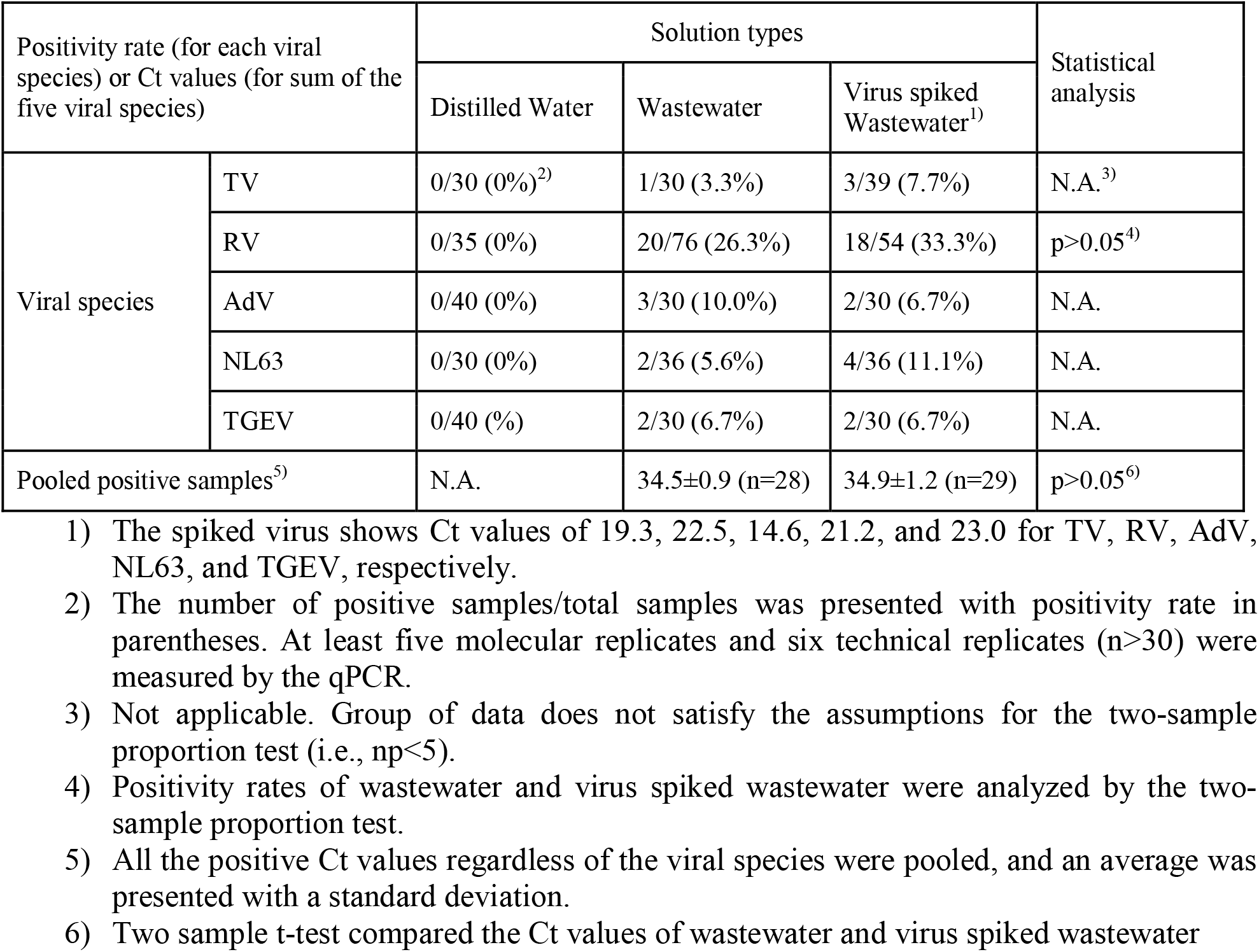
Specificity tests for the PGM-MBs method.

**Fig. 1.**
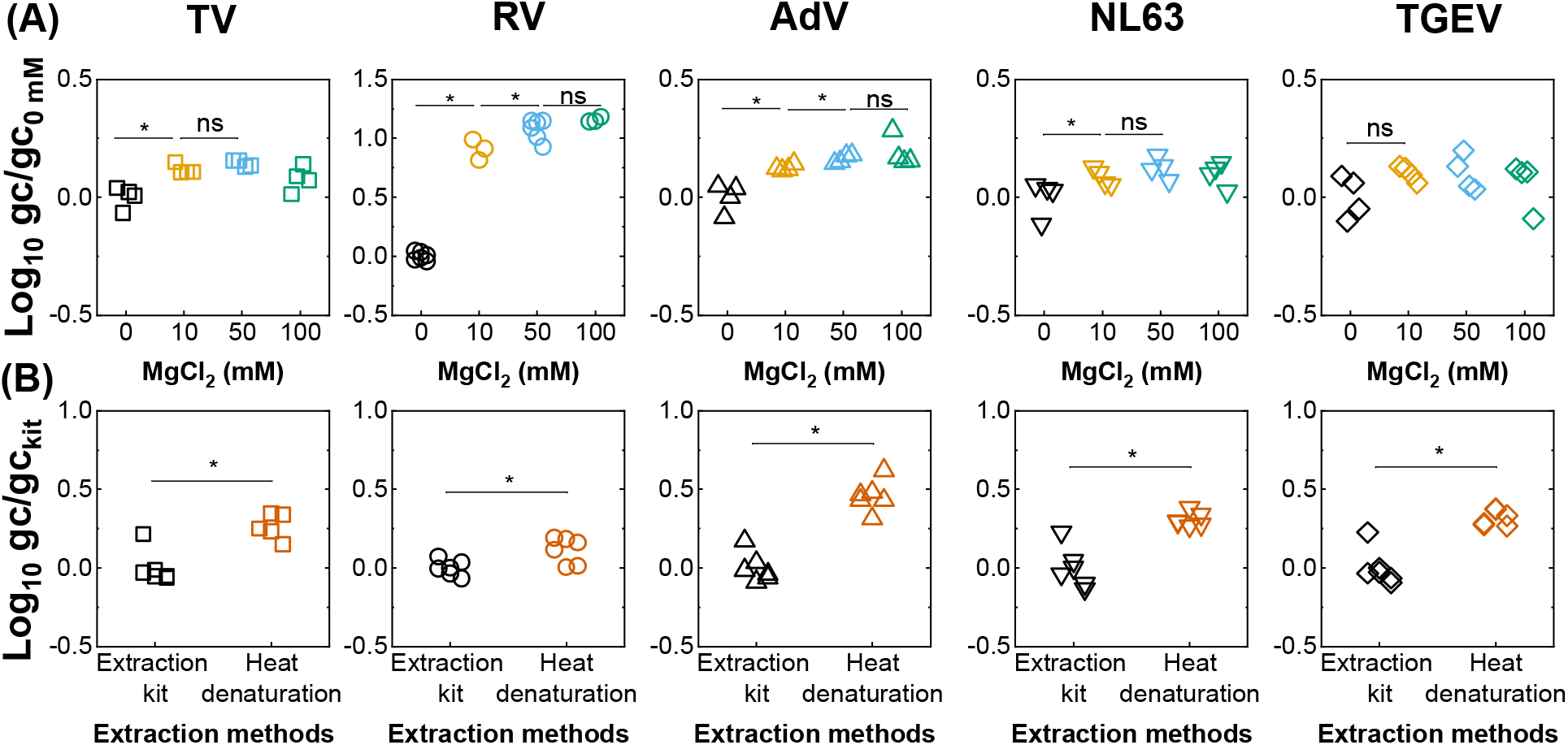
**(A)** Determination of the optimum MgCl_2_ concentration. The y-axis indicates the recovered viral genomes at different MgCl_2_ concentrations divided by the average number of viral genomes at 0 mM of MgCl_2_. **(B)** Comparison between results obtained with Viral RNA Mini Kit and with the heat denaturation method. The y-axis represents the recovered viral genomes divided by average number of viral genomes by extraction kit. Statistical analysis of two groups of results were conducted by a non-parametric test (Mann-Whitney Test; *: p<0.05 and ns: no significant difference).

**Fig. 2.**
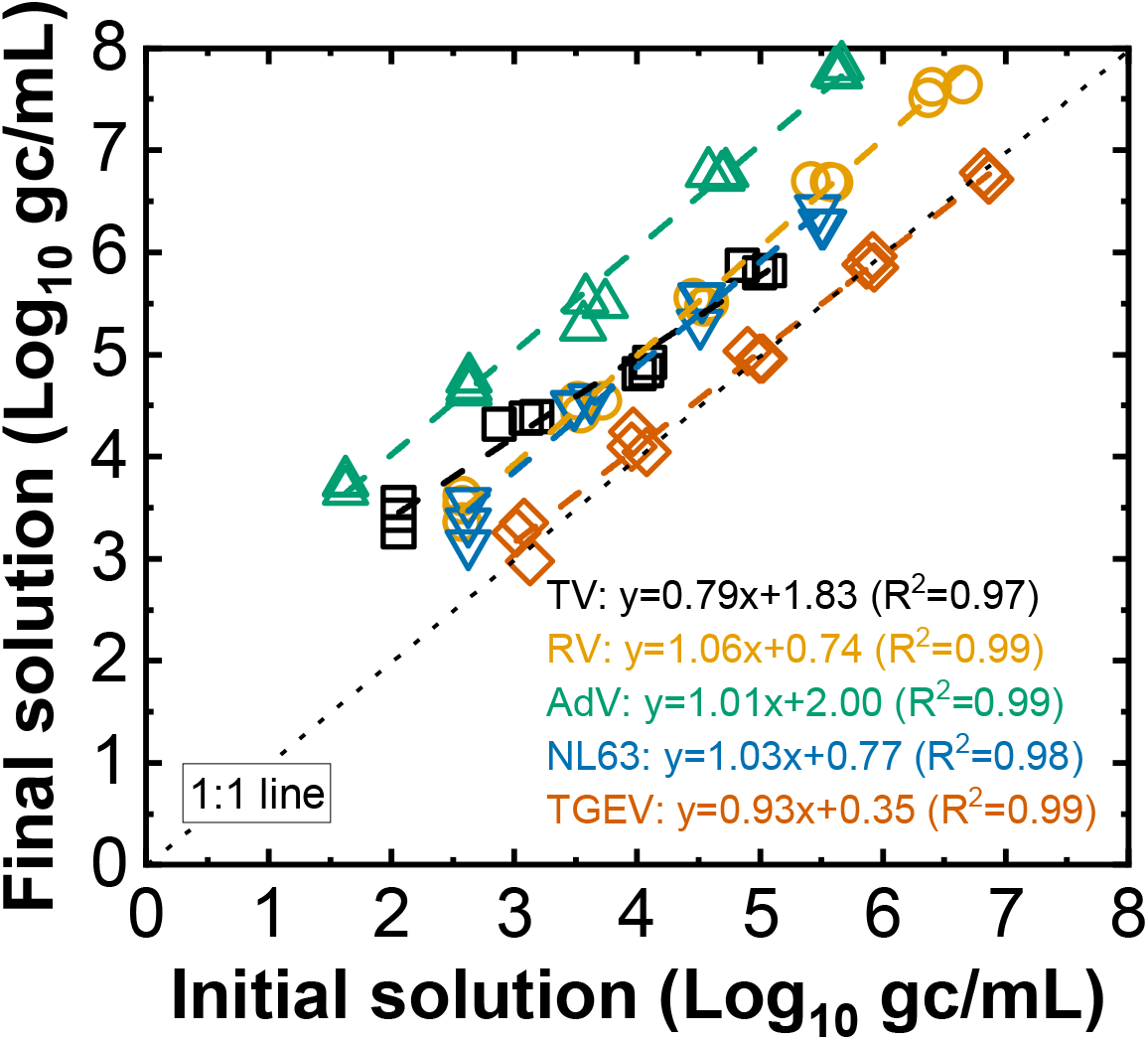
Calibration curves showing the relationship between genome concentrations of initial solutions and final solutions. Three biological replications were tested for each concentration.

**Fig. 3.**
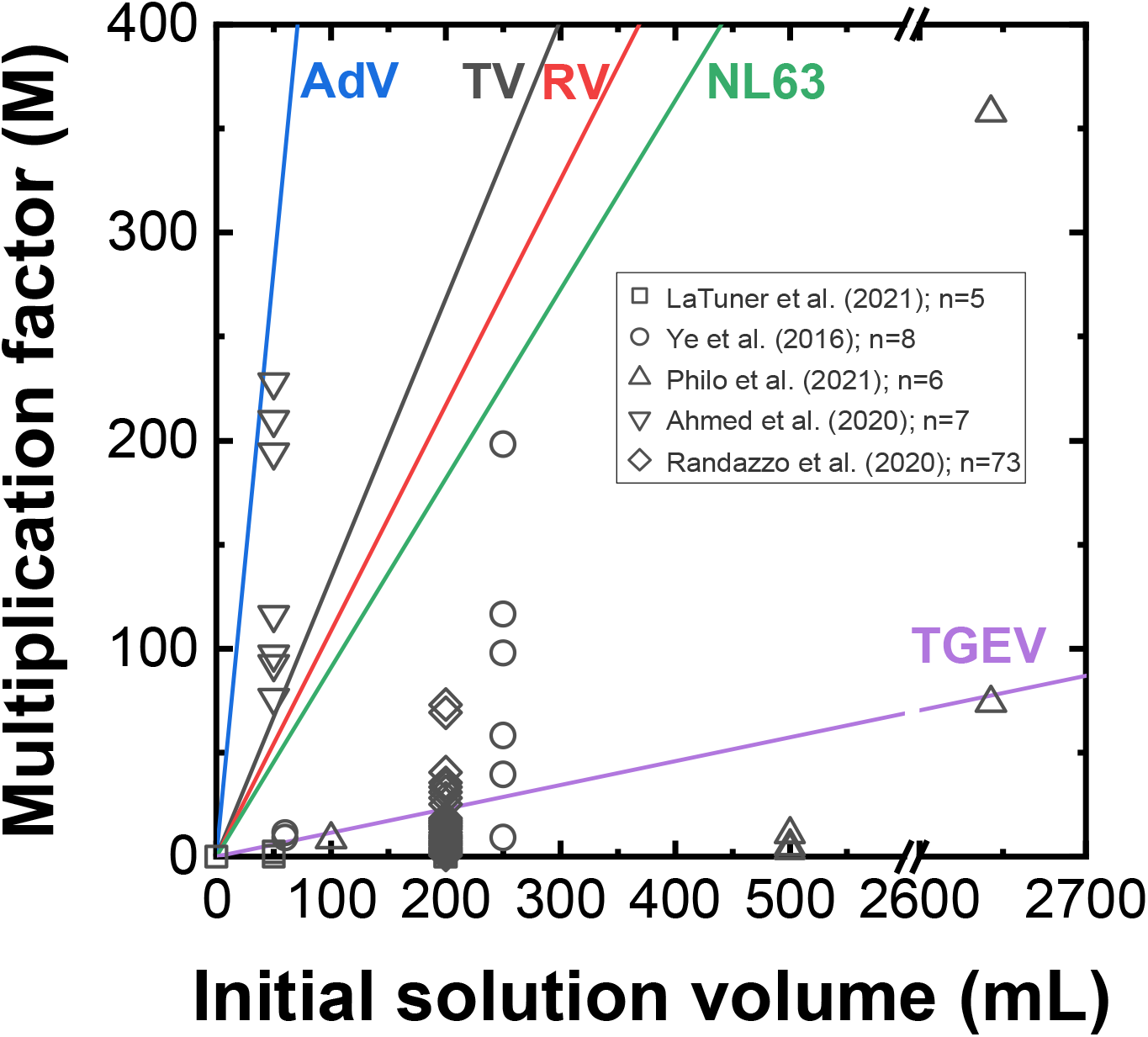
Comparisons of multiplication factors (MF) of the PGM-MBs method to those from other studies (Ahmed et al., 2020; LaTurner et al., 2021; Philo et al., 2021; Randazzo et al., 2020; Ye et al., 2016). Solid lines are extrapolated MF from experimental results with 10 mL of influent wastewater using Eq. 7. Different colors for the solid lines represent MF for different viral species determined by the PGM-MBs method. Open symbols are MFs calculated from 99 virus concentration experiments reported by 5 different studies. Different shapes for the open symbols indicate each reference.

**Fig. 4.**
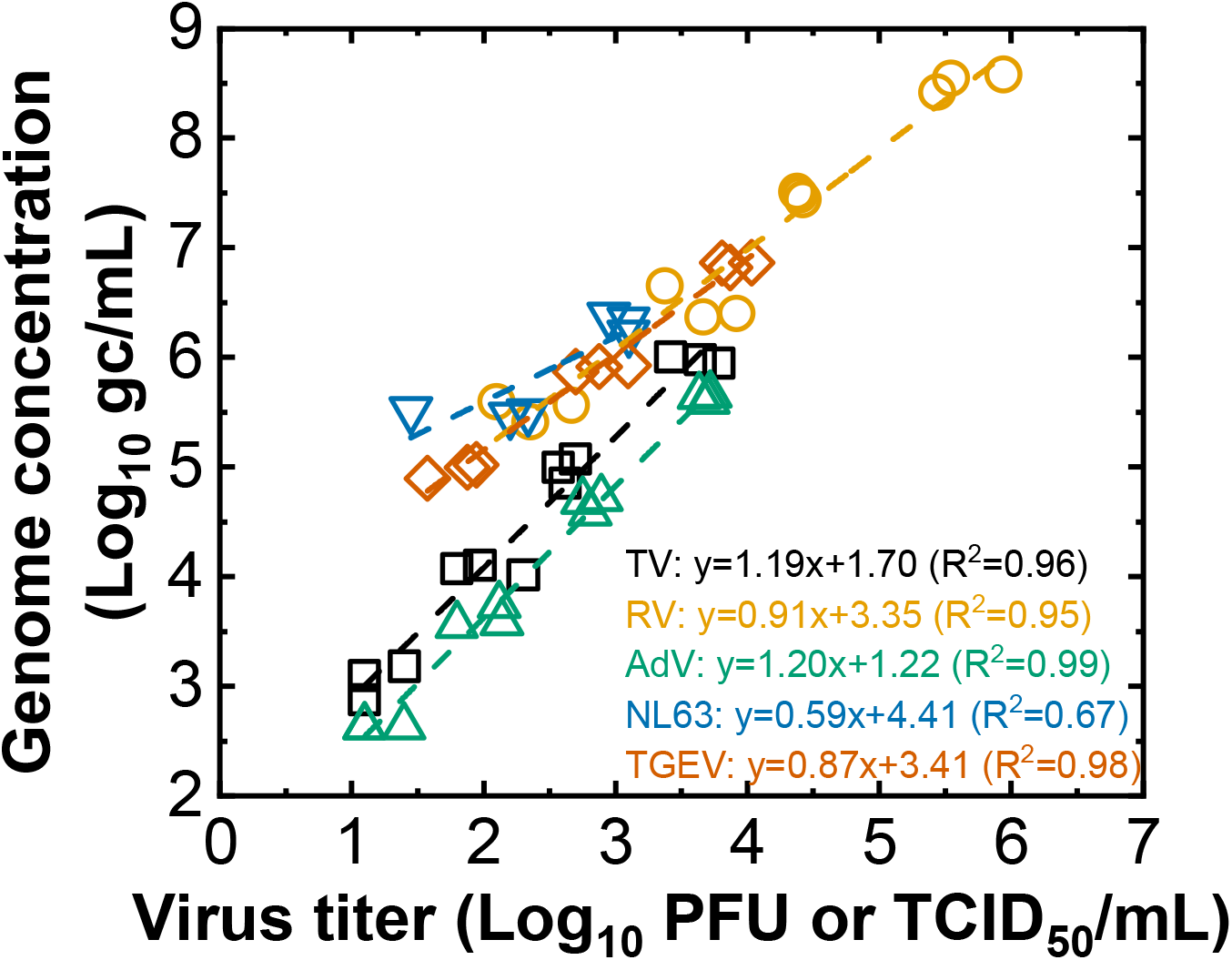
Calibration curves showing the relationship between infectious virus titers and genome concentrations of the virus-spiked wastewaters (n=3).

**Fig. 5.**
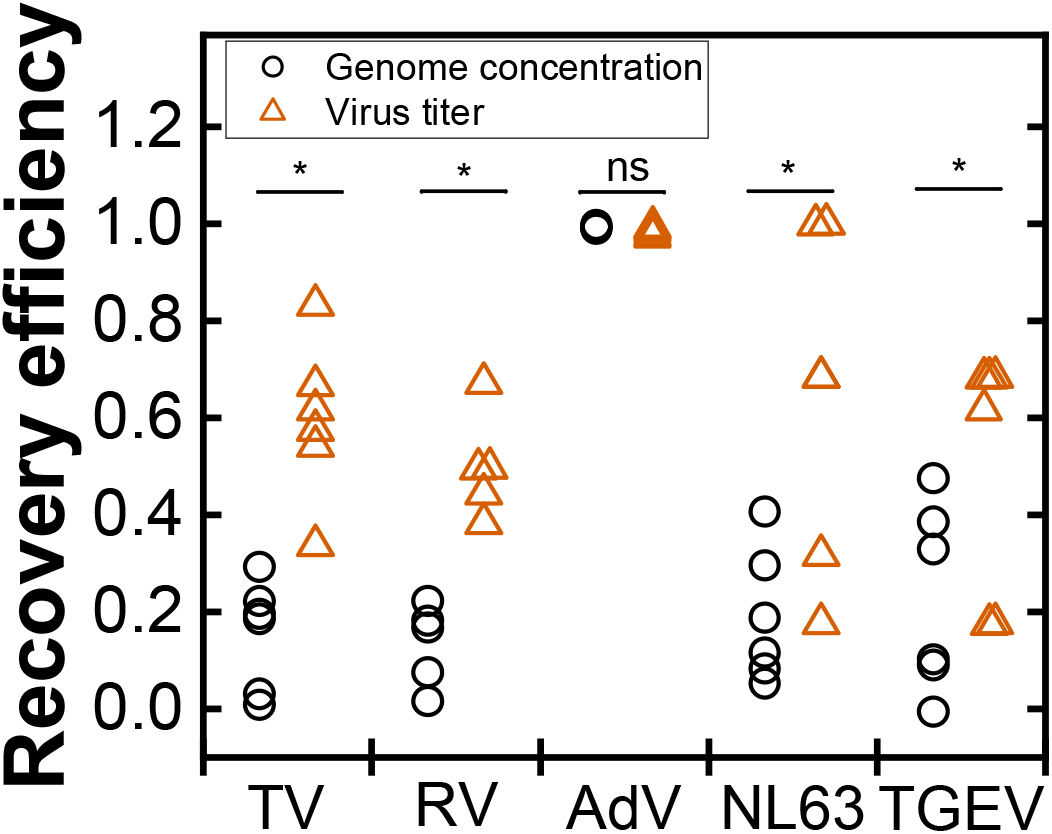
Recovery efficiency of the PGM-MBs method in terms of virus gene copy and virus titer. Statistical analyses were performed by a paired sample t-test (ns: no significant difference and *: p<0.05 and n=6).

**Fig. 6.**
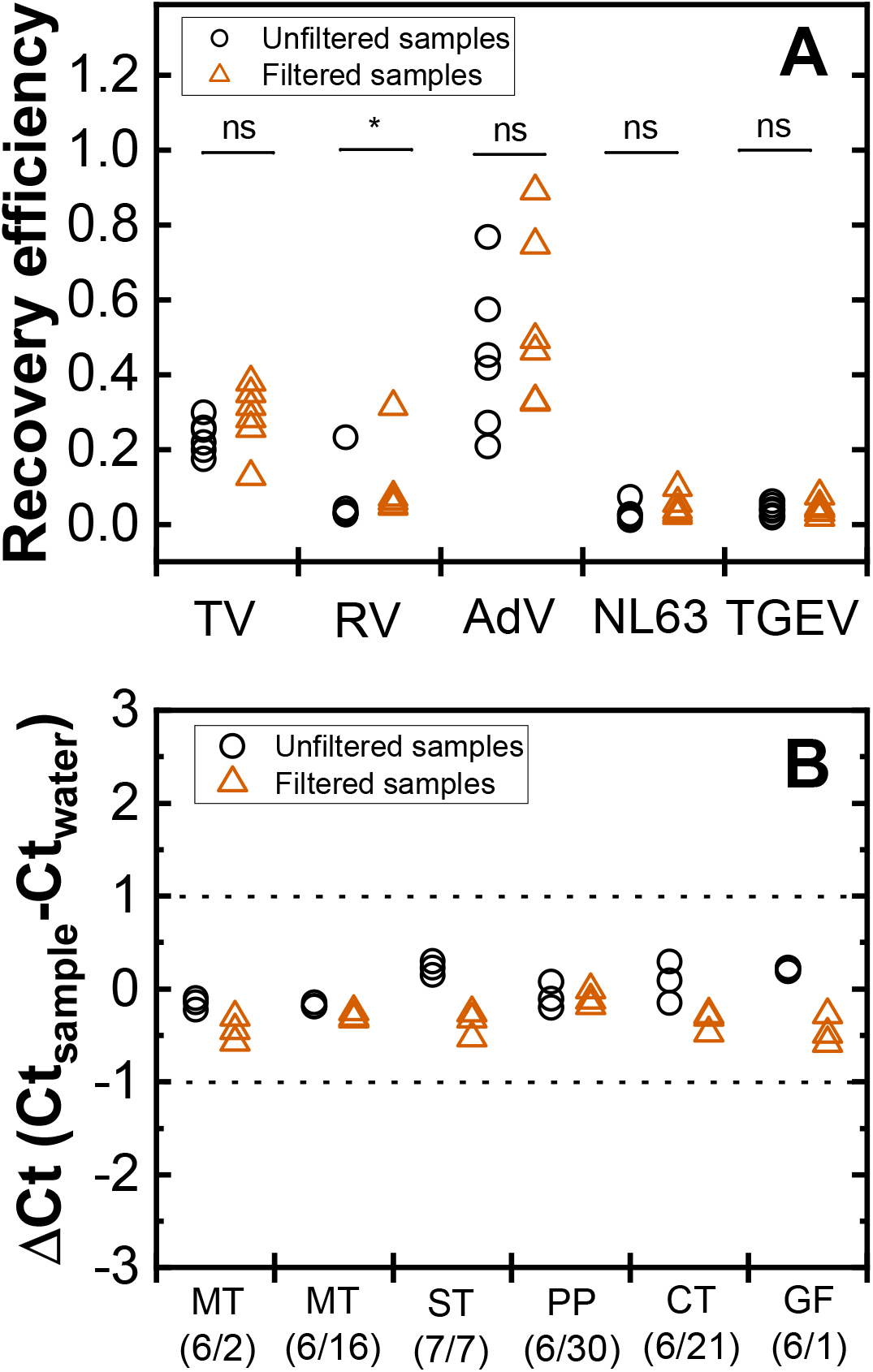
Impact of solid particles to **(A)** recovery efficiency and **(B)** tolerance to PCR inhibitors of the PGM-MBs method. **(A)** Each of five viral species were spiked to both filtered and unfiltered samples. Recovery efficiencies of both filtered and unfiltered samples for each viral species were compared by paired sample t-test (ns: no significant difference, *: p<0.05 and n=6). **(B)** Both filtered and unfiltered samples were subjected to the PGM-MBs method. TV genomes were spiked to final solutions of the PGM-MBs method and PCR inhibitor-free water (negative control). ΔCt values were calculated by subtracting Ct_water_ from Ct_sample_. One sample t-test was conducted twice with either a left or a right tail and all ΔCt values were within ±1 (p<0.05 and n=3).

## 3. Results

### 3.1. Optimization of the PGM-MBs method for efficient concentration of enteric viruses in wastewater

We tested MgCl_2_ concentration and genome extraction method to optimize the PGM-MBs method. MgCl_2_ is known to reduce repulsive electrical double layer force of small particles such as viruses in liquid (Gorrepati et al., 2010; Gutierrez and Nguyen, 2012), so it has been used to improve the performance of adsorption-based virus concentration methods such as electronegative membrane filtration method (Ahmed et al., 2020; LaTurner et al., 2021; Lu et al., 2020). We varied MgCl_2_ concentrations of the influent wastewater from 0 mM to 100 mM to find the optimum concentration for virus-PGM-MB interactions. **Fig. 1A** shows the amount of viruses recovered by PGM-MBs when different MgCl_2_ concentrations were used. With the exception of TGEV, MgCl_2_ increased the amount of virus binding to PGM-MBs. For example, the binding efficiency to PGM-MBs significantly increased for TV or NL63 as MgCl_2_ concentration increased until 10 mM (Mann-Whitney test, p<0.05). The binding efficiencies of RV or AdV to PGM-MBs became insignificantly different at 50 and 100 mM MgCl_2_ (Mann-Whitney test, p<0.05). Therefore, 50 mM was determined as the optimal concentration of MgCl_2_ for the PGM-MBs method.

Second, we established a heat denaturation method to release genomes from viruses bound to PGM-MBs as an alternative to using nucleic acid extraction kits. This method only requires the addition of proteinase K followed by heating up at 95□ for 10 minutes. **Fig. 1B** presents the results obtained from two experiments, in which the PGM-MBs method was applied to the spiked wastewater samples and the retrieved PGM-MBs were subjected to either an extraction kit (Viral RNA Mini Kit, Qiagen) or the heat denaturation method. The RNA extraction kit resulted in a significantly lower genome copy number than the heat denaturation method (Mann-Whitney Test p<0.05). Based on this comparsion, the alternative nucleic extraction mthod using Proteinase K and heat treatment can be recommended for PGM-MBs method.

### 3.2. Evaluation of the PGM-MBs method with spiked viruses

Genome concentrations of the spiked viruses in the initial wastewater samples are plotted on the X axis of **Fig. 2**. On the Y axis, we plotted the genome concentrations of the corresponding final solutions after applying the PGM-MBs method. Linear correlations were obtained for all cases (R^2^ >0.97) as shown in **Fig. 2**. Based on these linear correlations, we determined recovery efficiency (RE), concentration factor (CF), multiplication factor (MF), and LOQ_PGM-MBs_ as shown in Eqs 2-4.

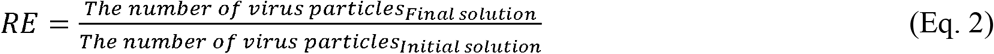

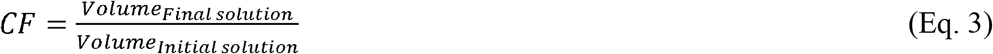

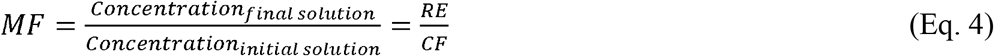

We summarized the average and standard deviation values for RE, CF, and MF obtained for each viral species in **Table 1**. We found that the PGM-MBs method showed different performances depending on testing viral species. For example, the REs ranged from 0.013 (TGEV) to 0.640 (AdV). The wide range of REs with different testing viruses were also reported by previous studies (Uchida et al., 2007; Ye et al., 2016). Since CF values were maintained throughout the experiments to be 0.01 (i.e., 0.1 mL of final volume and 10 mL of initial volume), MF values also showed a wide range from 1.3 (TGEV) to 64 (AdV). The MF values showed that AdV could be concentrated 64 times, while TGEV was concentrated by 1.3 times. Regardless of the viral species, the PGM-MBs method was able to concentrate enteric viruses from 10 mL of wastewater.

LOQ is defined by the lowest genome concentrations that fulfill the following two conditions: 1) None of the replicates (e.g., nine qPCR samples from three biological replicates and three technical replicates) is undetermined by qPCR or RT-qPCR analysis, and 2) coefficient of variation for all replicates is less than 25% (Forootan et al., 2017; Kralik and Ricchi, 2017). In this study, LOQ_F_ is LOQ of final solution obtained by the PGM-MBs method. On the other hand, the LOQ_PGM-MB_ is the genome concentrations of initial solution whose final concentrations after being concentrated by the PGM-MBs method becomes the LOQ_F_. By definition, LOQ_PGM-MBs_ is the lowest genome concentration of initial solution from **Fig. 2. Table 1** shows the LOQ of the PGM-MBs method for each viral species ranging from 10^4.63^ to 10^6.08^ gc/L. These LOQ values were comparable to the previously reported LOQs determined by 36 different methods for SARS-CoV-2 surveillance (Pecson et al., 2021).

We compared the MF values obtained by PGM-MBs method and conventional virus concentration methods applied in previous studies (Ahmed et al., 2020; LaTurner et al., 2021; Philo et al., 2021; Randazzo et al., 2020; Ye et al., 2016). Since the previous publications used different initial volumes, we calculated the MF values of PGM-MBs, which would be obtaind with wastewater samples used previously. For this calculation, we derived the following Eqs 5-7. Since LOQ_PGM-MBs_ is one of the initial concentrations at a specific condition where its final concentration is the LOQ_F_, we can derive Eq. 5 from Eqs. 2-4.

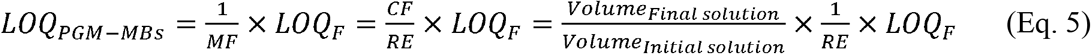

Eq. 5 shows that LOQ_PGM-MBs_ are inversely proportional to *Volume*_*Initial solution*_, so the LOQ_PGM-MBs_ will be lowered as the volume of the initial solution increases if the other variables such as *Volume*_*Final solution*_, *LOQ*_*F*_, and *RE* remain constant as *Volume*_*Initial solution*_ changes. Therefore, we can reasonably assume that *Volume*_*Final solution*_ and *LOQ*_*F*_ will be maintained if the same virus concentration method and quantification instruments are used. We also assumed RE is maintained when the volume ratio of the PGM-MBs to the initial solution is fixed to 10 µL to 10 mL. Therefore, we can derive Eqs. 6-7.

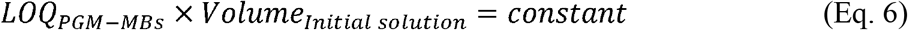

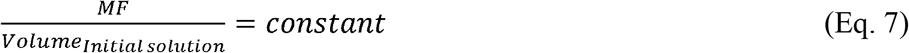

Eqs. 6-7 show LOQ_PGM-MBs_ and MF are functions of *Volume*_*Initial solution*_, and thus we can calculate LOQ_PGM-MBs_ and MF at different initial volumes such as 50, 200, and 1000 mL with LOQ_PGM-MBs_ and MF values that were experimentally determined with the 10 mL of the initial solution (**Table S4**).

We calculated the MF values of the PGM-MBs method with different initial volumes by Eq. 7. These MF values are presented as the solid lines in **Fig. 3**. Open symbols represent the MF of conventaionl virus concentration methods reported by previous studies (Ahmed et al., 2020; LaTurner et al., 2021; Philo et al., 2021; Randazzo et al., 2020; Ye et al., 2016). These previously reported MF values vary depending on virus concentration methods (direct extraction, electronegative membrane, PEG, ultrafiltration, ultracentrifugation, and skimmed milk etc.) and target viral species (BCoV, MHV, OC43, MgV, MS2, T3, and Phi6) (**Table S5**). Also, MF values will be also affected by water matrices that were not reflected in this comparison. Nevertheless, we found the PGM-MBs method presented higher MFs compared to the conventional counterparts in general. Specifically, MF values of PGM-MBs for TGEV (the least effective testing virus by the PGM-MBs method) were higher than 77 out of 99 MFs by conventional virus concentration methods.

### 3.3. Concentrating infectious viruses by the PGM-MBs method

The quantification of infectious virus titers in the environment are essential to evaluate accurately the risk posed to human health by viral pathogens (Haas et al., 2014). Thus, we designed experiments that evaluated the ability of PGM-MBs to concentrate infectious enteric viruses. The first experiment assessed if lab-grown viruses (TV, RV, AdV, NL63, and TGEV) spiked in wastewater could determine the environmentally relevant concentrations of these viruses. We serially diluted the lab-grown viruses to the filtered influent wastewater and determined the genome concentrations and infectious virus titers. The slopes in **Fig. 4** indicate the ratio of genome concentrations to infectious virus titers in the virus-spiked wastewater. As shown in **Fig. 4**, the genome concentrations are linearly correlated with the infectious virus titers for each virus with slopes ranging from 0.87 to 1.20 (R^2^>0.95) except for NL63 where only two serial dilutions were measured because the initial NL63 titer was lower (about 10^3^ TCID/mL) than the other species. This linear correlation is used to estimate the infectious virus titers at the environment with genome concentrations.

Next, we hypothesize that PGM-MBs have an intrinsic affinity to viruses with intact receptor-binding proteins that better represent infectious viruses than genome concentrations. This hypothesis is tested by experiments in which either genome concentrations or virus titers were used to determine RE. We assumed the PGM-MBs has an affinity to infectious virus particles if REs determined by virus titers are higher than those by genome concentrations. **Fig. 5** shows that RE calculated by virus titers (RE_titer_) are significantly higher than those by genome concentration (RE_genome_) for the tested viral species (Paired sample t-test, p<0.05) except for AdV where both RE_titer_ and RE_genome_ were close to 1 (Paired sample t-test, p>0.05). These results support our hypothesis that PGM-MBs have a higher affinity for infectious virus particles because the receptor-based approach excludes viruses whose receptor-binding proteins are deficient. The affinity to infectous virus particles is a unique feature of this PGM-MBs method considering the fact that conventional virus concentration methods such as ultracentrifugation, ultrafiltration, PEG, and electronegative filtration showed rather lower RE_genome_ compared to RE_titer_ (Rusiñol et al., 2020). Therefore, the recovered virus genomes by the PGM-MBs better represents the risk stemming from the enteric viruses in the environment than those determined by conventional counterparts.

### 3.4. Performance of the PGM-MBs method for unfiltered environmental samples

In the experiments described in 3.3, we removed bacterial cells that may interfere with the plaque assay by filtering the samples with 0.22 μm filters. Therefore, we designed another experiments to confirm that the PGM-MBs method works for environmental samples without the filtration process. We determined RE for each of the five viruses from the Filtered and Unfiltered sewage samples (**Fig. 6A)**. The paired sample t-test results showed that the differences between the Filtered and the Unfiltered samples were not significant different except for RV (p>0.05). This finding support that the presence of solid particles in the sewage samples did not significantly affected performances of the PGM-MBs method. We also tested the impact of solid particles as PCR inhibitors. **Fig. 6B** presented that ΔCt values for TV of the final solutions, which were obtained from either Filtered or Unfiltered sewage samples, were less than 1 (one sample t-test, p<0.05). This finding means that neglible inhibition impact, as suggested previously (Gibson et al., 2012; Wu et al., 2018). Thus, the PGM-MBs method can minimize the effect of PCR inhibitors regardless of the existence of solid particles in the unfiltered environmental samples.

### 3.5. Evaluation of the PGM-MBs method for PCR inhibitors in environmental samples

We designed experiments to test if the PGM-MBs method can reduce the PCR inhibitors from the environmental samples. PCR inhibition was evaluated by comparing the ΔCt values for 21 different samples (**Fig. 7**). Out of the 21 samples, 19 samples have ΔCt obtained by either the kit or PGM-MBs method is smaller than 1, so both the extraction kit and the PGM-MBs method were capable of eliminating PCR inhibitors for these 19 samples. For these 19 samples, there was no significant difference between ΔCt values of the two methods (paired sample t-test; p>0.05). On the other hand, both methods will require dilutions to avoid PCR inhibitors in the lagoon sample and samples spiked with coal-based humic acid. However, additional 5-fold and 2-fold dilution were enough for the PGM-MBs method to reduce PCR inhibitors remaining in the lagoon and humic acid samples, respectively. In contrast, those extra dilutions were not enough for the extraction kit to reduce the inhibition. Our findings agree with a previous study showing the PGM-MBs were less sensitive to the PCR inhibitors from fresh herbs and leafy vegetables than using PEG method and commercial genome extraction kit (Suresh et al., 2019). Maher et al. (2001) also demonstrated magnetic bead-based purification successfully eliminates PCR inhibitors in the airborne environment. Therefore, we concluded that the PGM-MBs method is as tolerant to PCR inhibitors in environmental samples as a commercial kit.

**Fig. 7.**
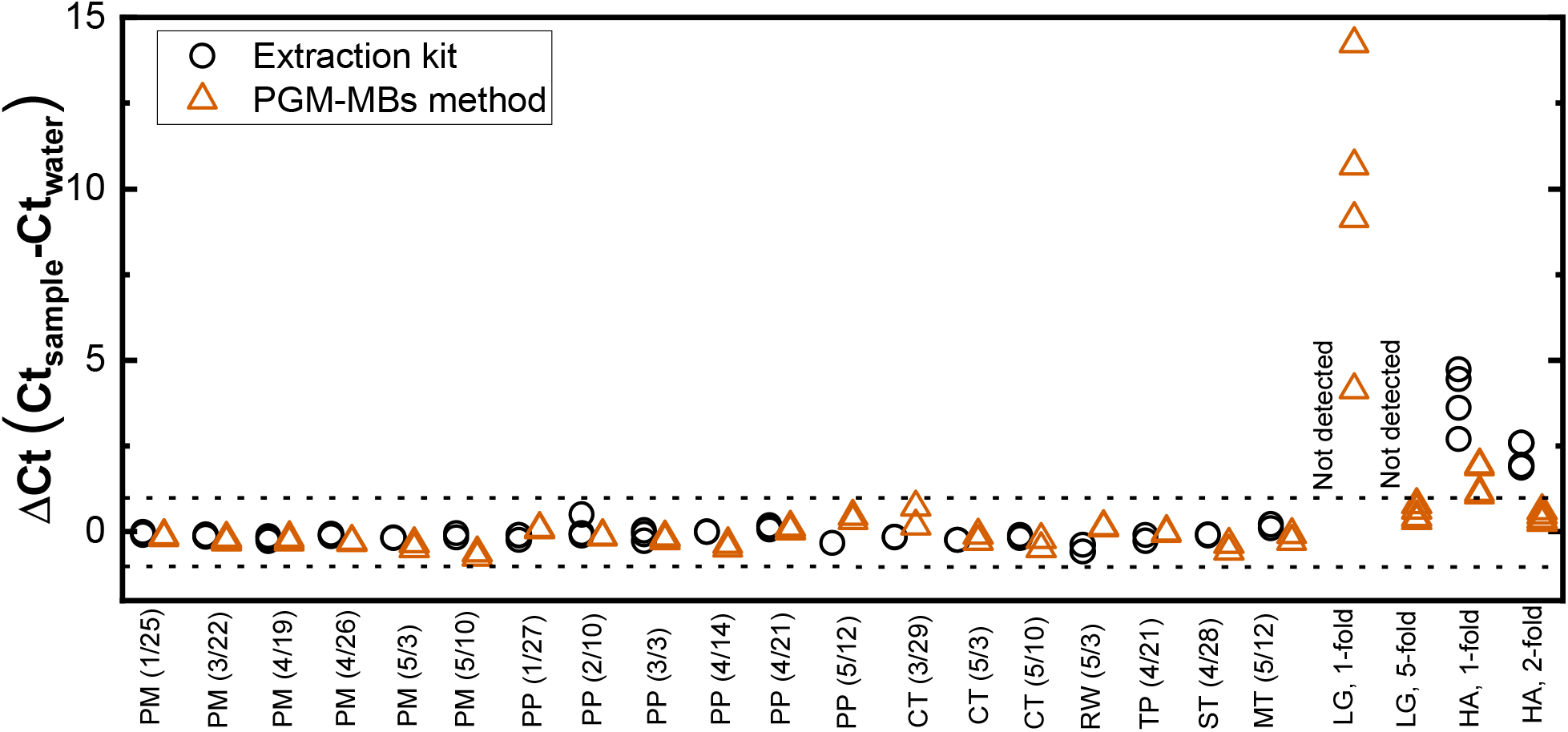
Evaluation of a commercial genome extraction kit (Viral RNA Mini Kit, Qiagen) and the PGM-MBs method in reducing PCR inhibitors in 20 environmental samples (different sampling locations or dates) and 20 mg/L of humic acid dissolved water. Tulane virus was spiked to those 21 different samples that were treated by either the extraction kit or the PGM-MBs method as well as molecular biology grade water (PCR inhibitor negative control). Lagoon and humic acid samples were further diluted 5-, and 2-fold, respectively.

### 3.6. Evaluation of PGM-MBs method for specificity

Porcine gastric mucin consists of different types of receptors, so various viral species are expected to bound when the PGM-MBs are applied to environmental samples (Larsson et al., 2009; McGuckin et al., 2011). The numbers of targeted virions attached to the PGM-MBs may be smaller than other virions which may also attach to the PGM-MBs. The attachment of these untargeted viral species could result in false-positive amplifications (Jaroenlak et al., 2016; Schrader et al., 2012; Tamariz et al., 2006). Therefore, we examined if the introduction of various viruses to the PGM-MBs causes false positives for the genome of our interest.

First, qPCR analysis of all 175 samples of PGM-MBs in molecular biology grade water showed no detected level of genomes for RV, AdV, NL63, and TGEV, indicating that viral genomes did not originate from the PGM-MBs (**Table 2**). Second, analysis of PGM-MBs in influent wastewater showed a few positive samples for TV, AdV, NL63, and TGEV, but 20 cases out of 76 (33%) for RV (**Table 2**). Because the primers used for our porcine RV targeted the VP1 gene of rotavirus group A, the detection of human RV in the wastewater is possible. *In Silico* analysis using Basic Local Alignment Search Tool (BLAST) indeed confirmed that the forward and reverse primers match 100% to human RVs (GenBank ID:LC389885.1 and ID:JQ715640.1, respectively). Human RV has been found in wastewater regardless of seasons (Atabakhsh et al., 2019; Ibrahim et al., 2016). Nevertheless, the proportion of RV positive samples from the influent wastewater were not significantly different from those from the virus-spiked wastewater (two sample proportion test; p>0.05) (**Table 2**). Ct values of pooled positive samples also did not show significant difference between the influent wastewater and the virus-spiked wastewater (two sample t-test; p>0.05). Considering the outweighing amount of the spiking viruses (Ct values ranging from 14 to 23) over the detected Ct values (about 35), we concluded that different types of viruses collected from the wastewater by the PGM-MBs method would not significantly affect the detection of target viruses.

### 3.7. Application of the PGM-MBs method for SARS-CoV-2 surveillance

We applied the electronegative membrane filtration method and the PGM-MBs methods in parallel for the SARS-CoV-2 surveillance with local sewages. The electronegative membrane filtration method represented conventional virus concentration methods here because of its high recovery efficiency (Ahmed et al., 2020; LaTurner et al., 2021; Lu et al., 2020). Seven different sewage samples, which tested positive for SARS-CoV-2 by either the filtration or the PGM-MBs method, were used for the comparison. To ensure qualities of the virus concentration processes, we spiked NL63, which recognizes the same receptors (ACE2) as SARS-CoV-2 (Rawat et al., 2021), to the sewage samples as an internal control. All the data reported showed higher than 1.0% of NL63 recovery efficiency. We found that NL63 recovery efficiencies for the filtration and the PGM-MBs method were normally distributed (Shapiro-Wilk test, p>0.05) in wide ranges from 1.4 to 18.6% (**Fig. S4**). The wide range of recovery efficiencies were also reported elsewhere (Randazzo et al., 2020), and this is probably because the two methods basically collect viral genomes through virus adsorption to their media (i.e., electronegative membrane filter and the PGM-MBs), which depends on the water characteristics (pH, ionic strength, or competing substances) (Gutierrez and Nguyen, 2012). The NL63 recovery efficiencies by the electronegative membrane filtration and the PGM-MBs method were not significantly different (paired sample t-test, p>0.05). We measured the N1 gene of SARS-CoV-2 (Nalla et al., 2020), and presented three technical replicates of Ct values, instead of concentrations, because most of the samples showed N1 gene concentrations below LOQ (**Table 3**). The Ct values for N1 gene were not significantly different between the two methods (paired sample t-test p>0.05). Therefore, we concluded the PGM-MBs method can be applied for monitoring SARS-CoV-2 in wastewater.

**Table 3.** Application of the PGM-MBs method for wastewater-based SARS-CoV-2 surveillance

| Sample<sup>1)</sup> | NL63 recovery (%) (multiplication factor) |  | Ct values for N1 gene |  |  |  |  |  |
| --- | --- | --- | --- | --- | --- | --- | --- | --- |
|  | Electronegative membrane filtration | PGM-MBs | Electronegative membrane filtration |  |  | PGM-MBs |  |  |
| RW (6/1) | 2.5 (21) | 6.1 (30) | 32.8<sup>1)</sup> | 33.2 | 34.2 | 33.9 | 33.1 | 33.4 |
| WS (6/1) | 2.0 (17) | 7.5 (38) | 36.7 | 35.0 | 36.3 | 36.4 | 34.9 | 36.4 |
| MT (6/9) | 1.7 (14) | 1.4 (7) | 34.1 | 33.6 | 34.0 | 37.2 | 36.5 | 36.4 |
| PM (6/13) | 9.7 (81) | 13.4 (67) | 35.9 | 37.8 | U<sup>2)</sup> | 35.9 | 36.0 | 37.0 |
| PM (6/21) | 12.9(107) | 18.6 (93) | 36.2 | 38.0 | U | 34.9 | 36.9 | 36.6 |
| ST (6/2) | 7.4 (64) | 3.7 (19) | 44.1 | U | U | 38.2 | U | U |
| ST (7/7) | 5.5 (46) | 3.9 (19) | 37.6 | 35.6 | U | 36.9 | 36.8 | U |

1) Numbers in paranthesis indicate collected month/date.
2) Whichever shaded data set between electronegative membrane filtration and PGM-MBs for each sample indicates lower average Ct values.
3) U stands for undetermined samples until 45 PCR cycles.

## 4. Discussion

### 4.1. The PGM-MBs method proves suitability for wastewater-based epidemiology

High throughput became an important factor for successful WBE because a fast turnaround for analyzing virus concentration of wastewater is critical (Betancourt et al., 2021; Zhu et al., 2021). The adaptability to high throughput instruments, low price, and a short operational time should be considered to evaluate virus concentration methods. Using magnetic beads and heat denaturation for collecting viruses and extracting viral genomes, respectively, allow the PGM-MBs method to satisfy those three requirements. Karthikeyan et al. (2021) demonstrated that a magnetic-bead-based approach could be implemented for an automated nucleic acid purification system (Thermo Fisher Scientific, USA), which enabled high throughput analysis (96 samples per run). In addition, using the PGM-MBs will further lower the cost for WBE. For example, consumables for the production of the 10 µL PGM-MBs (10 mL wastewater sample analysis) cost 0.413 USD (**Table S6**). Essential materials for this method, including magnetic beads, mucin, magnets, and proteinase K are not proprietary. Also, the entire process for the PGM-MBs method only takes less than 3 hours, including concentrating viruses (30 minutes), extracting genomes (10 minutes), and quantifying viral genomes (90 minutes), which is much shorter than conventional virus concentration methods (Cervantes-Avilés et al., 2021). For example, ultrafiltration (Haramoto et al., 2020), skimmed milk flocculation (Guerrero-Latorre et al., 2020), and PEG precipitation (La Rosa et al., 2020b) took 5.2, 9.6, and 12.8 hours only for a virus concentration step, respectively. Taken together, the PGM-MBs method is expected to be scaled up for high throughput analysis.

### 4.2. Performance of PGM-MBs method is comparable to or better than conventional virus concentration methods

We made two comparisons between PGM-MBs and conventional counterparts to evaluate the performances in concentrating viruses from environmental samples. First, we systematically characterize the performances of PGM-MBs method with five model viruses and wastewater. Because there are various virus concentration methods, we could not test all these conventional methods in the same experimental conditions as the experiments for the PGM-MBs method. Instead, we compared MF of the PGM-MBs method to those reported by previous studies. Note that we chose MF over LOQ for the comparison because LOQ depends on instruments (e.g., ddPCR versus qPCR) (Falzone et al., 2020; Park et al., 2021) and qPCR methods (e.g., SYBR versus Taqman method) (Fuchs Wightman et al., 2021) while MF focuses only on the increase in viral genome concentrations by virus concentration methods. Thus, although we could compare the PGM-MBs method with various experiments using different virus concentrations (**Fig. 3**), we could not directly compare the results on the same experimental conditions. To compensate for this limitation, we applied the PGM-MBs method and electronegative membrane filtration method, which is one of the most efficient virus conventional methods (Ahmed et al., 2020; LaTurner et al., 2021; Lu et al., 2020) for SARS-CoV-2 concentration under the same experimental conditions (**Table 3**). Given the findings from **Fig. 3** and **Table 3**, we concluded that the PGM-MBs method is comparable to or better than conventional methods to concentrate various enteric viruses, including SARS-CoV-2 from environmental samples.

## 5. Conclusions

This study first introduced a novel approach to concentrate enteric viruses from the environment using porcine gastric mucin-conjugated magnetic beads (PGM-MBs). This novel method is simple, fast, affordable, and comparable with conventional methods. The optimized PGM-MBs method takes less than 3 hours from virus concentration to genome quantification and costs less than 0.5 USD for 10 mL volume of sample without expensive instruments such as an ultracentrifuge. We systematically demonstrated that the performance of PGM-MBs method is comparable to or better than conventional methods to concentrate various enteric viruses, including SARS-CoV-2, from environmental samples. We also discovered that the PGM-MBs method is robust to environmental samples, which features the existence of different viral species and PCR inhibitors. Taken all together, we concluded that the PGM-MBs method can readily be used for urgent SARS-CoV-2 surveillance to cope with the current COVID-19 pandemic or monitoring other enteric viruses for better public health management.

## Data Availability

All data produced in the present study are available upon reasonable request to the authors

## Acknowledgement

This project is funded by the Grainger College of Engineering and the JUMP-ARCHES program of OSF Healthcare in conjunction with the University of Illinois. The Human coronavirus NL63 strain (NR-470) was obtained through BEI Resources, NIAID. We thank Mr. Bruce Rabe at Urbana Ȧ Champaign Sanitary District for providing us with influent wastewater. We also acknowledge Bill Brown for sampling site selection, Hayden Wennerdahl, Kip Stevenson, Dr. Laura Keefer and Dr. Schmidt for sampling deployment, and Yuqing Mao, Aijia Zhou, Matthew Robert Loula, Aashna Patra, Kristin Joy Anderson, Mikayla Diedrick, Hubert Lyu, Hamza Elmahi Mohamed, Jad R Karajeh, Runsen Ning, Rui Fu, Kate O’Brien for sewage sampling and processing.

## Reference

Afolayan, O.T., Webb, C.C., Cannon, J.L., 2016. Evaluation of a Porcine Gastric Mucin and RNase A Assay for the Discrimination of Infectious and Non-infectious GI.1 and GII.4 Norovirus Following Thermal, Ethanol, or Levulinic Acid Plus Sodium Dodecyl Sulfate Treatments. Food Environ. Virol. 8, 70–78. https://doi.org/10.1007/s12560-015-9219-z

Ahmed, W., Bertsch, P.M., Bivins, A., Bibby, K., Farkas, K., Gathercole, A., Haramoto, E., Gyawali, P., Korajkic, A., McMinn, B.R., Mueller, J.F., Simpson, S.L., Smith, W.J.M., Symonds, E.M., Thomas, K. V., Verhagen, R., Kitajima, M., 2020. Comparison of virus concentration methods for the RT-qPCR-based recovery of murine hepatitis virus, a surrogate for SARS-CoV-2 from untreated wastewater. Sci. Total Environ. 739, 139960. https://doi.org/10.1016/J.SCITOTENV.2020.139960

Ahmed, W., Tscharke, B., Bertsch, P.M., Bibby, K., Bivins, A., Choi, P., Clarke, L., Dwyer, J., Edson, J., Nguyen, T.M.H., O’Brien, J.W., Simpson, S.L., Sherman, P., Thomas, K. V., Verhagen, R., Zaugg, J., Mueller, J.F., 2021. SARS-CoV-2 RNA monitoring in wastewater as a potential early warning system for COVID-19 transmission in the community: A temporal case study. Sci. Total Environ. 761, 144216. https://doi.org/10.1016/J.SCITOTENV.2020.144216

Albinana-Gimenez, N., Clemente-Casares, P., Bofill-Mas, S., Hundesa, A., Ribas, F., Girones, R., 2006. Distribution of Human Polyoma-viruses, Adenoviruses, and Hepatitis E Virus in the Environment and in a Drinking-Water Treatment Plant†. Environ. Sci. Technol. 40, 7416–7422. https://doi.org/10.1021/ES060343I

Araud, E., Shisler, J.L., Nguyen, T.H., 2018. Inactivation Mechanisms of Human and Animal Rotaviruses by Solar UVA and Visible Light. Environ. Sci. Technol. 52, 5682–5690. https://doi.org/10.1021/acs.est.7b06562

Atabakhsh, P., Kargar, M., Doosti, A., 2019. Molecular detection and genotyping of group A rotavirus in two wastewater treatment plants, Iran. Brazilian J. Microbiol. 2019 511 51, 197–203. https://doi.org/10.1007/S42770-019-00131-0

Betancourt, W.Q., Schmitz, B.W., Innes, G.K., Prasek, S.M., Pogreba Brown, K.M., Stark, E.R., Foster, A.R., Sprissler, R.S., Harris, D.T., Sherchan, S.P., Gerba, C.P., Pepper, I.L., 2021. COVID-19 containment on a college campus via wastewater-based epidemiology, targeted clinical testing and an intervention. Sci. Total Environ. 779, 146408. https://doi.org/10.1016/J.SCITOTENV.2021.146408

Binder, A.M., Biggs, H.M., Haynes, A.K., Chommanard, C., Lu, X., Erdman, D.D., Watson, J.T., Gerber, S.I., 2017. Human Adenovirus Surveillance — United States, 2003–2016. MMWR. Morb. Mortal. Wkly. Rep. 66, 1039. https://doi.org/10.15585/MMWR.MM6639A2

Bustin, S.A., Benes, V., Garson, J.A., Hellemans, J., Huggett, J., Kubista, M., Mueller, R., Nolan, T., Pfaffl, M.W., Shipley, G.L., Vandesompele, J., Wittwer, C.T., 2009. The MIQE guidelines: Minimum information for publication of quantitative real-time PCR experiments. Clin. Chem. 55, 611–622. https://doi.org/10.1373/clinchem.2008.112797

Cervantes-Avilés, P., Moreno-Andrade, I., Carrillo-Reyes, J., 2021. Approaches applied to detect SARS-CoV-2 in wastewater and perspectives post-COVID-19. J. Water Process Eng. 40, 101947. https://doi.org/10.1016/J.JWPE.2021.101947

Falzone, L., Musso, N., Gattuso, G., Bongiorno, D., Palermo, C.I., Scalia, G., Libra, M., Stefani, S., 2020. Sensitivity assessment of droplet digital PCR for SARS-CoV-2 detection. Int. J. Mol. Med. 46, 957–964. https://doi.org/10.3892/IJMM.2020.4673/HTML

Fielding, B.C., 2011. Human coronavirus NL63: a clinically important virus? Future Microbiol. 6, 153–159. https://doi.org/10.2217/FMB.10.166

Forootan, A., Sjöback, R., Björkman, J., Sjögreen, B., Linz, L., Kubista, M., 2017. Methods to determine limit of detection and limit of quantification in quantitative real-time PCR (qPCR). Biomol. Detect. Quantif. 12, 1. https://doi.org/10.1016/J.BDQ.2017.04.001

Fuchs Wightman, F., Godoy Herz, M.A., Muñoz, J.C., Stigliano, J.N., Bragado, L., Moreno, N.N., Palavecino, M., Servi, L., Cabrerizo, G., Clemente, J., Avaro, M., Pontoriero, A., Benedetti, E., Baumeister, E., Rudolf, F., Remes Lenicov, F., Garcia, C., Buggiano, V., Kornblihtt, A.R., Srebrow, A., de la Mata, M., Muñoz, M.J., Schor, I.E., Petrillo, E., 2021. A DNA intercalating dye-based RT-qPCR alternative to diagnose SARS-CoV-2. RNA Biol. https://doi.org/10.1080/15476286.2021.1926648/SUPPL_FILE/KRNB_A_1926648_SM7808.ZIP

Fuzawa, M., Araud, E., Li, J., Shisler, J.L., Nguyen, T.H., 2019. Free Chlorine Disinfection Mechanisms of Rotaviruses and Human Norovirus Surrogate Tulane Virus Attached to Fresh Produce Surfaces. Environ. Sci. Technol. 53, 11999–12006. https://doi.org/10.1021/acs.est.9b03461

Gibas, C., Lambirth, K., Mittal, N., Juel, M.A.I., Barua, V.B., Roppolo Brazell, L., Hinton, K., Lontai, J., Stark, N., Young, I., Quach, C., Russ, M., Kauer, J., Nicolosi, B., Chen, D., Akella, S., Tang, W., Schlueter, J., Munir, M., 2021. Implementing building-level SARS-CoV-2 wastewater surveillance on a university campus. Sci. Total Environ. 782, 146749. https://doi.org/10.1016/J.SCITOTENV.2021.146749

Gibson, K.E., Schwab, K.J., Spencer, S.K., Borchardt, M.A., 2012. Measuring and mitigating inhibition during quantitative real time PCR analysis of viral nucleic acid extracts from large-volume environmental water samples. Water Res. 46, 4281–4291. https://doi.org/10.1016/J.WATRES.2012.04.030

Gorrepati, E.A., Wongthahan, P., Raha, S., Fogler, H.S., 2010. Silica precipitation in acidic solutions: Mechanism, pH effect, and salt effect. Langmuir 26, 10467–10474. https://doi.org/10.1021/LA904685X/SUPPL_FILE/LA904685X_SI_001.PDF

Guerrero-Latorre, L., Ballesteros, I., Villacrés-Granda, I., Granda, M.G., Freire-Paspuel, B., Ríos-Touma, B., 2020. SARS-CoV-2 in river water: Implications in low sanitation countries. Sci. Total Environ. 743, 140832. https://doi.org/10.1016/J.SCITOTENV.2020.140832

Gutierrez, L., Nguyen, T.H., 2012. Interactions between rotavirus and Suwannee River organic matter: Aggregation, deposition, and adhesion force measurement. Environ. Sci. Technol. 46, 8705–8713. https://doi.org/10.1021/ES301336U/SUPPL_FILE/ES301336U_SI_001.PDF

Haas, C.N., Rose, J.B., Gerba, C.P., 2014. Quantitative Microbial Risk Assessment. John Wiley & Sons.

Haramoto, E., Kitajima, M., Hata, A., Torrey, J.R., Masago, Y., Sano, D., Katayama, H., 2018. A review on recent progress in the detection methods and prevalence of human enteric viruses in water. Water Res. 135, 168–186. https://doi.org/10.1016/J.WATRES.2018.02.004

Haramoto, E., Malla, B., Thakali, O., Kitajima, M., 2020. First environmental surveillance for the presence of SARS-CoV-2 RNA in wastewater and river water in Japan. Sci. Total Environ. 737, 140405. https://doi.org/10.1016/J.SCITOTENV.2020.140405

Harris-Lovett, S., Nelson, K.L., Beamer, P., Bischel, H.N., Bivins, A., Bruder, A., Butler, C., Camenisch, T.D., De Long, S.K., Karthikeyan, S., Larsen, D.A., Meierdiercks, K., Mouser, P.J., Pagsuyoin, S., Prasek, S.M., Radniecki, T.S., Ram, J.L., Keith Roper, D., Safford, H., Sherchan, S.P., Shuster, W., Stalder, T., Wheeler, R.T., Korfmacher, K.S., 2021. Wastewater Surveillance for SARS-CoV-2 on College Campuses: Initial Efforts, Lessons Learned, and Research Needs. Int. J. Environ. Res. Public Health 18. https://doi.org/10.3390/IJERPH18094455

Hart, O.E., Halden, R.U., 2020. Computational analysis of SARS-CoV-2/COVID-19 surveillance by wastewater-based epidemiology locally and globally: Feasibility, economy, opportunities and challenges. Sci. Total Environ. 730, 138875. https://doi.org/10.1016/J.SCITOTENV.2020.138875

Hu, Zhiliang, Song, C., Xu, C., Jin, G., Chen, Y., Xu, X., Ma, H., Chen, W., Lin, Y., Zheng, Y., Wang, J., Hu, Zhibin, Yi, Y., Shen, H., 2020. Clinical characteristics of 24 asymptomatic infections with COVID-19 screened among close contacts in Nanjing, China. Sci. China Life Sci. 63, 706–711. https://doi.org/10.1007/s11427-020-1661-4

Ibrahim, C., Cherif, N., Hammami, S., Pothier, P., Hassen, A., 2016. Quantification and Genotyping of Rotavirus A within Two Wastewater Treatment Processes. CLEAN–Soil, Air, Water 44, 393–401. https://doi.org/10.1002/CLEN.201400588

Jafferali, M.H., Khatami, K., Atasoy, M., Birgersson, M., Williams, C., Cetecioglu, Z., 2021. Benchmarking virus concentration methods for quantification of SARS-CoV-2 in raw wastewater. Sci. Total Environ. 755, 142939. https://doi.org/10.1016/J.SCITOTENV.2020.142939

Jaroenlak, P., Sanguanrut, P., Williams, B.A.P., Stentiford, G.D., Flegel, T.W., Sritunyalucksana, K., Itsathitphaisarn, O., 2016. A Nested PCR Assay to Avoid False Positive Detection of the Microsporidian Enterocytozoon hepatopenaei (EHP) in Environmental Samples in Shrimp Farms. PLoS One 11, e0166320. https://doi.org/10.1371/JOURNAL.PONE.0166320

Kang, G., 2017. Viral Diarrhea. Int. Encycl. Public Heal. 360. https://doi.org/10.1016/B978-0-12-803678-5.00486-0

Kapikian, A.Z., 1996. Overview of viral gastroenteritis. Arch. Virol. Suppl. 1996, 7–19. https://doi.org/10.1007/978-3-7091-6553-9_2

Karthikeyan, S., Ronquillo, N., Belda-Ferre, P., Alvarado, D., Javidi, T., Longhurst, C.A., Knight, R., 2021. High-Throughput Wastewater SARS-CoV-2 Detection Enables Forecasting of Community Infection Dynamics in San Diego County. mSystems 6. https://doi.org/10.1128/MSYSTEMS.00045-21/SUPPL_FILE/MSYSTEMS.00045-21-ST003.DOCX

Katayama, H., Shimasaki, A., Ohgaki, S., 2002. Development of a Virus Concentration Method and Its Application to Detection of Enterovirus and Norwalk Virus from Coastal Seawater. Appl. Environ. Microbiol. 68, 1033–1039. https://doi.org/10.1128/AEM.68.3.1033-1039.2002

Kralik, P., Ricchi, M., 2017. A basic guide to real time PCR in microbial diagnostics: Definitions, parameters, and everything. Front. Microbiol. 8, 108. https://doi.org/10.3389/FMICB.2017.00108/BIBTEX

La Rosa, G., Bonadonna, L., Lucentini, L., Kenmoe, S., Suffredini, E., 2020a. Coronavirus in water environments: Occurrence, persistence and concentration methods - A scoping review. Water Res. 179, 115899. https://doi.org/10.1016/J.WATRES.2020.115899

La Rosa, G., Iaconelli, M., Mancini, P., Bonanno Ferraro, G., Veneri, C., Bonadonna, L., Lucentini, L., Suffredini, E., 2020b. First detection of SARS-CoV-2 in untreated wastewaters in Italy. Sci. Total Environ. 736, 139652. https://doi.org/10.1016/J.SCITOTENV.2020.139652

Larsson, J.M.H., Karlsson, H., Sjövall, H., Hansson, G.C., 2009. A complex, but uniform O-glycosylation of the human MUC2 mucin from colonic biopsies analyzed by nanoLC/MSn. Glycobiology 19, 756–766. https://doi.org/10.1093/GLYCOB/CWP048

LaTurner, Z.W., Zong, D.M., Kalvapalle, P., Gamas, K.R., Terwilliger, A., Crosby, T., Ali, P., Avadhanula, V., Santos, H.H., Weesner, K., Hopkins, L., Piedra, P.A., Maresso, A.W., Stadler, L.B., 2021. Evaluating recovery, cost, and throughput of different concentration methods for SARS-CoV-2 wastewater-based epidemiology. Water Res. 197, 117043. https://doi.org/10.1016/J.WATRES.2021.117043

Lodder, W.J., De Roda Husman, A.M., 2005. Presence of noroviruses and other enteric viruses in sewage and surface waters in The Netherlands. Appl. Environ. Microbiol. 71, 1453–1461. https://doi.org/10.1128/AEM.71.3.1453-1461.2005

Lu, D., Huang, Z., Luo, J., Zhang, X., Sha, S., 2020. Primary concentration–The critical step in implementing the wastewater based epidemiology for the COVID-19 pandemic: A mini-review. Sci. Total Environ. 747, 141245. https://doi.org/10.1016/j.scitotenv.2020.141245

Maher, N., Dillon, H.K., Vermund, S.H., Unnasch, T.R., 2001. Magnetic bead capture eliminates PCR inhibitors in samples collected from the airborne environment, permitting detection of Pneumocystis carinii DNA. Appl. Environ. Microbiol. 67, 449–452. https://doi.org/10.1128/AEM.67.1.449-452.2001/ASSET/2BB3A5B0-1602-400E-A0C3-C32CFC5AFC72/ASSETS/GRAPHIC/AM0110760003.JPEG

McGuckin, M.A., Lindén, S.K., Sutton, P., Florin, T.H., 2011. Mucin dynamics and enteric pathogens. Nat. Rev. Microbiol. 2011 94 9, 265–278. https://doi.org/10.1038/nrmicro2538

Nalla, A.K., Casto, A.M., Huang, M.-L.W., Perchetti, G.A., Sampoleo, R., Shrestha, L., Wei, Y., Zhu, H., Jerome, K.R., Greninger, A.L., 2020. Comparative Performance of SARS-CoV-2 Detection Assays Using Seven Different Primer-Probe Sets and One Assay Kit. https://doi.org/10.2807/1560-7917.ES.2020

Nemudryi, A., Nemudraia, A., Wiegand, T., Surya, K., Buyukyoruk, M., Cicha, C., Vanderwood, K.K., Wilkinson, R., Wiedenheft, B., 2020. Temporal Detection and Phylogenetic Assessment of SARS-CoV-2 in Municipal Wastewater. Cell Reports Med. 1, 100098. https://doi.org/10.1016/J.XCRM.2020.100098

Oh, C., Sun, P.P., Araud, E., Nguyen, T.H., 2020. Mechanism and efficacy of virus inactivation by a microplasma UV lamp generating monochromatic UV irradiation at 222 nm. Water Res. 186. https://doi.org/10.1016/j.watres.2020.116386

Panchal, D., Prakash, O., Bobde, P., Pal, S., 2021. SARS-CoV-2: sewage surveillance as an early warning system and challenges in developing countries. Environ. Sci. Pollut. Res. 2021 2818 28, 22221–22240. https://doi.org/10.1007/S11356-021-13170-8

Park, C., Lee, Jina, ul Hassan, Z., Ku, K.B., Kim, S.J., Kim, H.G., Park, E.C., Park, G.S., Park, Daeui, Baek, S.H., Park, Dongju, Lee, Jihye, Jeon, S., Kim, Seungtaek, Lee, C.S., Yoo, H.M., Kim, Seil, 2021. Comparison of Digital PCR and Quantitative PCR with Various SARS-CoV-2 Primer-Probe Sets. J. Microbiol. Biotechnol. 31, 358–367. https://doi.org/10.4014/JMB.2009.09006

Pecson, B.M., Darby, E., Haas, C.N., Amha, Y.M., Bartolo, M., Danielson, R., Dearborn, Y., Di Giovanni, G., Ferguson, C., Fevig, S., Gaddis, E., Gray, D., Lukasik, G., Mull, B., Olivas, L., Olivieri, A., Qu, Y., SARS-CoV-2 Interlaboratory Consortium, 2021. Reproducibility and sensitivity of 36 methods to quantify the SARS-CoV-2 genetic signal in raw wastewater: findings from an interlaboratory methods evaluation in the U.S. Environ. Sci. Water Res. Technol. https://doi.org/10.1039/d0ew00946f

Philo, S.E., Keim, E.K., Swanstrom, R., Ong, A.Q.W., Burnor, E.A., Kossik, A.L., Harrison, J.C., Demeke, B.A., Zhou, N.A., Beck, N.K., Shirai, J.H., Meschke, J.S., 2021. A comparison of SARS-CoV-2 wastewater concentration methods for environmental surveillance. Sci. Total Environ. 760, 144215. https://doi.org/10.1016/J.SCITOTENV.2020.144215

Polo, D., Quintela-Baluja, M., Corbishley, A., Jones, D.L., Singer, A.C., Graham, D.W., Romalde, J.L., 2020. Making waves: Wastewater-based epidemiology for COVID-19–approaches and challenges for surveillance and prediction. Water Res. 186, 116404. https://doi.org/10.1016/J.WATRES.2020.116404

Randazzo, W., Truchado, P., Cuevas-Ferrando, E., Simón, P., Allende, A., Sánchez, G., 2020. SARS-CoV-2 RNA in wastewater anticipated COVID-19 occurrence in a low prevalence area. Water Res. 181, 115942. https://doi.org/10.1016/J.WATRES.2020.115942

Rawat, P., Jemimah, S., Ponnuswamy, P.K., Gromiha, M.M., 2021. Why are ACE2 binding coronavirus strains SARS-CoV/SARS-CoV-2 wild and NL63 mild? Proteins Struct. Funct. Bioinforma. 89, 389–398. https://doi.org/10.1002/PROT.26024

Rusiñol, M., Martínez-Puchol, S., Forés, E., Itarte, M., Girones, R., Bofill-Mas, S., 2020. Concentration methods for the quantification of coronavirus and other potentially pandemic enveloped virus from wastewater. Curr. Opin. Environ. Sci. Heal. 17, 21–28. https://doi.org/10.1016/J.COESH.2020.08.002

Satyanarayana, M., 2020. Shortage of RNA extraction kits hampers efforts to ramp up COVID-19 coronavirus testing [WWW Document]. URL https://cen.acs.org/analytical-chemistry/diagnostics/Shortage-RNA-extraction-kits-hampers/98/web/2020/03 (accessed 11.23.21).

Schrader, C., Schielke, A., Ellerbroek, L., Johne, R., 2012. PCR inhibitors - occurrence, properties and removal. J. Appl. Microbiol. 113, 1014–1026. https://doi.org/10.1111/j.1365-2672.2012.05384.x

Sherchan, S.P., Shahin, S., Patel, J., Ward, L.M., Tandukar, S., Uprety, S., Schmitz, B.W., Ahmed, W., Simpson, S., Gyawali, P., 2021. Occurrence of SARS-CoV-2 RNA in Six Municipal Wastewater Treatment Plants at the Early Stage of COVID-19 Pandemic in The United States. Pathog. (Basel, Switzerland) 10. https://doi.org/10.3390/PATHOGENS10070798

Suresh, M., Harlow, J., Nasheri, N., 2019. Evaluation of porcine gastric mucin assay for detection and quantification of human norovirus in fresh herbs and leafy vegetables. Food Microbiol. 84, 103254. https://doi.org/10.1016/J.FM.2019.103254

Tamariz, J., Voynarovska, K., Prinz, M., Caragine, T., 2006. The Application of Ultraviolet Irradiation to Exogenous Sources of DNA in Plasticware and Water for the Amplification of Low Copy Number DNA. J. Forensic Sci. 51, 790–794. https://doi.org/10.1111/J.1556-4029.2006.00172.X

Uchida, E., Kogi, M., Oshizawa, T., Furuta, B., Satoh, K., Iwata, A., Murata, M., Hikata, M., Yamaguchi, T., 2007. Optimization of the virus concentration method using polyethyleneimine-conjugated magnetic beads and its application to the detection of human hepatitis A, B and C viruses. J. Virol. Methods 143, 95–103. https://doi.org/10.1016/J.JVIROMET.2007.02.014

Walker, D.I., Cross, L.J., Stapleton, T.A., Jenkins, C.L., Lees, D.N., Lowther, J.A., 2019. Assessment of the Applicability of Capsid-Integrity Assays for Detecting Infectious Norovirus Inactivated by Heat or UV Irradiation. Food Environ. Virol. 11, 229–237. https://doi.org/10.1007/s12560-019-09390-4

Waruhiu, C., Ommeh, S., Obanda, V., Agwanda, B., Gakuya, F., Ge, X.Y., Yang, X. Lou, Wu, L.J., Zohaib, A., Hu, B., Shi, Z.L., 2017. Molecular detection of viruses in Kenyan bats and discovery of novel astroviruses, caliciviruses and rotaviruses. Virol. Sin. 2017 322 32, 101–114. https://doi.org/10.1007/S12250-016-3930-2

Wolfe, M.K., Archana, A., Catoe, D., Coffman, M.M., Dorevich, S., Graham, K.E., Kim, S., Grijalva, L.M., Roldan-Hernandez, L., Silverman, A.I., Sinnott-Armstrong, N., Vugia, D.J., Yu, A.T., Zambrana, W., Wigginton, K.R., Boehm, A.B., 2021. Scaling of SARS-CoV-2 RNA in Settled Solids from Multiple Wastewater Treatment Plants to Compare Incidence Rates of Laboratory-Confirmed COVID-19 in Their Sewersheds. Environ. Sci. Technol. Lett. 8, 398–404. https://doi.org/10.1021/ACS.ESTLETT.1C00184

Wu, Q., Kawano, K., Uehara, Y., Okuda, N., Hongo, M., Tsuji, S., Yamanaka, H., Minamoto, T., 2018. Environmental DNA reveals nonmigratory individuals of Palaemon paucidens overwintering in Lake Biwa shallow waters. Freshw. Sci. 37, 307–314. https://doi.org/10.1086/697542/ASSET/IMAGES/LARGE/FG3.JPEG

Ye, Y., Ellenberg, R.M., Graham, K.E., Wigginton, K.R., 2016. Survivability, Partitioning, and Recovery of Enveloped Viruses in Untreated Municipal Wastewater. Environ. Sci. Technol. 50, 5077–5085. https://doi.org/10.1021/ACS.EST.6B00876/SUPPL_FILE/ES6B00876_SI_001.PDF

Yu, G., Zhang, D., Guo, F., Tan, M., Jiang, X., Jiang, W., 2013. Cryo-EM Structure of a Novel Calicivirus, Tulane Virus. PLoS One 8, 59817. https://doi.org/10.1371/journal.pone.0059817

Zhu, Y., Oishi, W., Maruo, C., Saito, M., Chen, R., Kitajima, M., Sano, D., 2021. Early warning of COVID-19 via wastewater-based epidemiology: potential and bottlenecks. Sci. Total Environ. 767, 145124. https://doi.org/10.1016/J.SCITOTENV.2021.145124

