## Supplemental Figures and Tables for "A novel approach to concentrate human and animal viruses from wastewater using receptors-conjugated magnetic beads"

**Text S1. Preparation of porcine gastric mucin conjugated magnetic beads (PGM-MBs)**

The porcine gastric mucin conjugated magnetic beads (PGM-MBs) were produced based on our previous studies (Araud et al., 2018; Fuzawa et al., 2019; Oh et al., 2020). One milliliter of MagnaBind carboxyl-derivatized beads (Thermo Scientific, USA) was put into a 1.5 mL low adhesion centrifuge tube (USA Scientific). The magnetic beads were rinsed by vortexing it with clean PBS while a bead attractor (S1509S, New Biolabs Lab, USA) held the beads. This washing process was repeated three times. Ten milligrams of porcine gastric mucin (M1778, Sigma-Aldrich, USA) was dissolved into 1 mL of MES (28390, Thermo Scientific, USA). Ten milliliters of EDC solution (22980, Thermo Scientific, USA) was dissolved in another 1 mL MES. One milliliter of the mucin solution and 0.1 mL of the EDC solution were added to the washed magnetic beads. The mixture was shaken on the orbital shaker (Fisher Scientific, USA) at 400 rpm for 30 minutes, during which the porcine gastric mucin was conjugated to the magnetic beads. The PGM-MBs were separated by the attractor and the supernatant was replaced by the clean PBS. After that, the PGM-MBs solution was washed by vortexing for 10 seconds. The washing process was repeated three times. The PGM-MBs were resuspended in 1 mL PBS with 0.05% sodium azide and stored at 4℃ until use.

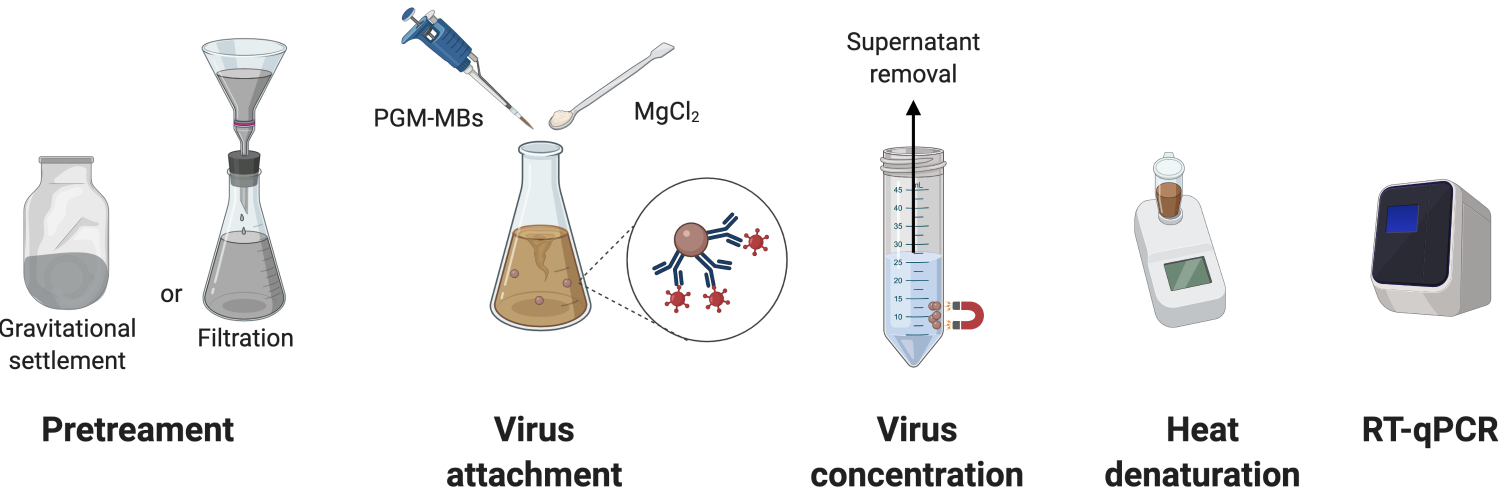

**Fig. S1.** A procedure for the PGM-MBs method.

**Table S1.** The checklist from MIQE guidelines and relevant information for this study

| **Item to check** | **Location** |
| --- | --- |
| 1. Experimental design | |
| Definition of experimental and control groups | Materials and Methods |
| Number within each group | Materials and Methods; Each figure |
| 2. Sample | |
| Description | Materials and Methods (Section 2.4) |
| Volume/mass of sample processed | Materials and Methods (each section) |
| Processing procedure | Materials and Methods (Section 2.4) |
| Sample storage conditions and duration | Materials and Methods (Section 2.4) |
| 3. Nucleic acid extraction | |
| Procedure and/or instrumentation | Materials and Methods (Section 2.2, SI Text 1, SI Table 2) |
| Name of kit and details of any modifications | Materials and Methods (Section 2.2, SI Text 1, SI Table 2) |
| Contamination assessment (DNA or RNA) | Materials and Methods (Section 2.2, SI Text 1, SI Table 2) |
| Nucleic acid quantification | Materials and Methods (Section 2.2, SI Text 1, SI Table 2) |
| Instrument and method | Materials and Methods (Section 2.2, SI Text 1, SI Table 2) |
| Inhibition testing (C_q_ dilutions, spike, or other) | Fig. 7 |
| 4. Reverse transcription | |
| Complete reaction conditions | Materials and Methods (Section 2.2, SI Text 1, SI Table 2) |
| Amount of RNA and reaction volume | Materials and Methods (Section 2.2, SI Text 1, SI Table 2) |
| Reverse transcriptase and concentration | Materials and Methods (Section 2.2, SI Text 1, SI Table 2) |
| Temperature and time | Materials and Methods (Section 2.2, SI Text 1, SI Table 2) |
| Manufacturer of reagents and catalogue numbers | Materials and Methods (Section 2.2, SI Text 1, SI Table 2) |
| 5. qPCR target information | |
| Gene symbol | SI Table 2 |
| Sequence accession number | SI Table 2 |
| Location of amplicon | SI Table 2 |
| Amplicon length | SI Table 2 |
| In silico specificity screen (BLAST, and so on) | SI Table 2 |
| 6. qPCR oligonucleotides | |
| Primer sequences | SI Table 2 |
| Manufacturer of oligonucleotides | SI Table 2 |
| 7. qPCR protocol | |
| Complete reaction conditions | Materials and Methods (Section 2.2, SI Text 1, SI Table 2) |
| Reaction volume and amount of DNA | Materials and Methods (Section 2.2, SI Text 1, SI Table 2) |
| Primer (probe) concentrations | Materials and Methods (Section 2.2, SI Text 1, SI Table 2) |
| Polymerase identity and concentration | Materials and Methods (Section 2.2, SI Text 1, SI Table 2) |
| Buffer/kit identity and manufacturer | Materials and Methods (Section 2.2, SI Text 1, SI Table 2) |
| Manufacturer of plates/tubes and catalog number | Materials and Methods (Section 2.2, SI Text 1, SI Table 2) |
| Complete thermocycling parameters | Materials and Methods (Section 2.2, SI Text 1, SI Table 2) |
| Manufacturer of qPCR instrument | Materials and Methods (Section 2.2, SI Text 1, SI Table 2) |
| 8. qPCR validation | |
| Specificity (gel, sequence, melt, or digest) | Materials and Methods (Section 2.2, SI Text 1, SI Table 2) |
| For SYBR Green I, C_q_ of the NTC | Materials and Methods (Section 2.2, SI Text 1, SI Table 2) |
| Calibration curves with slope and *y* intercept | Materials and Methods (Section 2.2, SI Text 1, SI Table 2) |
| PCR efficiency calculated from slope | Materials and Methods (Section 2.2, SI Text 1, SI Table 2) |
| *r*2 of calibration curve | Materials and Methods (Section 2.2, SI Text 1, SI Table 2) |
| Linear dynamic range | Materials and Methods (Section 2.2, SI Text 1, SI Table 2) |
| C_q_ variation at LOD | Materials and Methods (Section 2.2, SI Text 1, SI Table 2) |
| Evidence for LOD | Materials and Methods (Section 2.2, SI Text 1, SI Table 2) |
| 9. Data analysis | |
| qPCR analysis program (source, version) | Materials and Methods (Section 2.2, SI Text 1) |
| Method of C_q_ determination | Materials and Methods (Section 2.2, SI Text 1) |
| Outlier identification and disposition | Materials and Methods (Section 2.2, SI Text 1) |
| Results for NTCs | Materials and Methods (Section 2.2, SI Text 1) |
| Description of normalization method | Materials and Methods (Section 2.2, SI Text 1) |
| Number and concordance of biological replicates | Materials and Methods; each Figure |
| Number and stage of technical replicates | Materials and Methods (Section 2.2, SI Text 1) |
| Repeatability (intraassay variation) | Materials and Methods (Section 2.2, SI Text 1) |
| Statistical methods for results significance | Each figure |
| Software (source, version) | Materials and Methods |
| Data transparency | Raw data available upon request |

**Text S2. Viral nucleic acid extraction and quantification**

The SYBR-based RT-qPCR started with mixing the 3 μL of viral genome with 5 μL of 2 × iTaq universal SYBR green reaction mix, 0.125 μL of iScript reverse transcriptase from the iTaq universal SYBR green reaction mix (Bio-Rad Laboratories, USA), 0.3 μL of 10 μM forward primer for each virus, 0.3 μL of 10 μM reverse primer for each virus, and 1.275 μL of molecular biology grade water (Corning, NY, USA). The PCR cocktail for the one-step RT-qPCR was placed in 96-well plates (4306737, Applied Biosystems, USA) and analyzed by a qPCR system (QuantStudio 3, Thermo Fisher Scientific, USA). The thermocycle began with 10 minutes at 50℃ and 1 minute at 90℃ followed by 40 cycles of 30 seconds at 60℃ and 1 minute at 90℃. The Taqman-based RT-qPCR was initiated by mixing 5 μL of viral genome with 5 μL of Taqman Fast Virus 1-step Master Mix (4444432, Applied Biosystems, USA), 1.5 μL of primers/probe mixture for N1 gene (2019-nCoV RUO kit, Integrated DNA Technologies, USA), and 8.5 μL of water. The 20 μL of mixture was analyzed by the same qPCR system as used for the SYBR-based RT-qPCR, except for a different thermal cycle (5 minutes at 50℃, 20 seconds at 95℃ followed by 45 cycles of 3 seconds at 95℃ and 30 seconds at 55℃). In case of DNA viruses, the 2 µL of viral genome was mixed with 5.0 μL of PowerUp SYBR™ Green Master Mix (Applied Biosystems, CA, USA), 0.3 μL of 10 μM forward primer, 0.3 μL of 10 μM reverse primer, and 2.4 μL of the molecular biology grade water (Corning, NY, USA). The qPCR cocktail was also placed in the 96-well plates (4306737, Applied Biosystems, USA) and the qPCR was ran using a qPCR system (QuantStudio 3, Thermo Fisher Scientific, USA) with the following thermocycle: 10 minutes at 95°C; 40 cycles of 15 seconds at 95°C and 1 minute at 60°C; and temperature increase from 65°C to 95°C for melting curve analysis. PCR standard curves were obtained for every one-step RT-qPCR and qPCR analysis with 10-fold serial dilutions of synthetic DNA oligonucleotide (Integrated DNA technologies, USA) and PCR efficiencies for the one-step RT-qPCR was higher than 85% (R^2^>0.99) and those for the qPCR was higher than 90% (R^2^>0.99). The detailed information about the primers and synthetic DNA controls were summarized in **SI Table 2**.

**Table S2.** Summary of Primers and thermocycles information for qPCR analysis

| Assay type | Viral species | Primer name | Sequence^1)^ (5’-3’) | Position in the genome | Amplicon length (bp) | Reaction conditions |
| --- | --- | --- | --- | --- | --- | --- |
| One-step RT-qPCR | TV | TV-NSP1-F | GTGCGCATCCTTGAGACAAT | 879-899 | 133^2)^ | 50°C for 10 min and 95°C for 1 min, followed by 40 cycles of (95°C for 10 s, 60°C for 30 sec) |
|  |  | TV-NSP1-R | TTGGAGCCGGGTAGAAACAT | 991-1011 |  |  |
|  | RV | RV-VP1-F | ACGATAAGTATAATGCTGTAGAACGG | 253–279 | 109^3)^ |  |
|  |  | RV-VP1-R | CAGCTGATTGGAATAATTCAGAAGT | 285–361 |  |  |
|  | NL63 | NL63-ORF1a/1b-F | TGGAAGCAACGTTCTGTCGT | 243-262 | 137^4)^ |  |
|  |  | NL63-ORF1a/1b-R | ATAGGTGCGAACGGCTACAG | 360-379 |  |  |
|  | TGEV | TGEV-ORF1a-F | TTGTCTTCGGACACCAACTC | 71-90 | 101^5)^ |  |
|  |  | TGEV-ORF1b-R | GGAGTGATTGCCAAACTGAATG | 150-171 |  |  |
| qPCR | AdV | AdV-L5-F | TTACTAGCGGCGCTCTTAGC | 31394-31413 | 115^6)^ | 95°C for 10 min, 40 cycles of (95°C for 15 s, 60°C for 1 min) |
|  |  | AdV-L5-R | TGCAGGGCTAGCTTTCCATC | 31489-31508 |  |  |

1) Primer pair specificity was checked by the primer-blast tool (National Center of Biotechnology Information). Each pair of primers were blasted with their host organism (e.g., Rhesus monkey (txid:9544) for MA104 and MK2 cell lines; Sus scrofa (txid:9823) for ST cells; Homo sapiens (txid:9606) for A549 cells). We confirmed that our primers did not target any sequences of their host cells.
2) The sequence of standard sample for the NSP1 gene of TV (Integrated DNA technologies, USA): 5’-AGAATTGGACCGAATTTGGCACACACTCAGAATTTGGTGTGCGCATCCTTGAGACAATAACAGGCACAATACCCCCTTGGAAACCTCACCAGGAATCAATATCTGAAGTTCTGGACGACCTCACACACGGTAAAGTCCAAACAGGTGATGATGTTTCTACCCGGCTCCAAAGGTTGAGCGACACTATCAAAGATCTGAGTGTCATGGCTTGTGATCCCTCTGCACCGCCCGAAGTTGCGC-3’ (GenBank accession number: EU391643).
3) The sequence of standard sample for the VP1 gene of RV (Integrated DNA technologies, USA): 5’-GCATCCATATTATCGTATTCCTACGATAAGTATAATGCTGTAGAACGGAAGTTAGTTAATTATGCTAAAGGTAAACCATTAGAAGCAGATTTGACGGCGAATGAACTCGATTATGAAAATAATAAAATAACTTCTGAATTATTCCAATCAGCTGAAGAATATACTGATTCATTAATGGATCCTGCTATACTGACTTCATTATCATCTAATTTAAATGCAGTTATGTTTTGGTTGGAACG-3’ (GenBank accession number: MT025928.1)
4) The sequence of standard sample for the ORF1a/1b gene of NL63 (Integrated DNA technologies, USA): 5’-TTCTCAACTAAACGAAATTTTTCTAGTGCTGTCATTTGTTATGGCAGTCCTAGTGTAATTGAAATTTCGTCAAGTTTGTAAACTGGTTAGGCAAGTGTTGTATTTTCTGTGTCTAAGCACTGGTGATTCTGTTCACTAGTGCATACATTGATATTTAAGTGGTGTTCCGTCACTGCTTATTGTGGAAGCAACGTTCTGTCGTTGTGGAAACCAATAACTGCTAACCATGTTTTACAATCAAGTGACACTTGCTGTTGCAAGTGATTCGGAAATTTCAGGTTTTGGTTTTGCCATTCCTTCTGTAGCCGTTCGCACCTATAGCGAAGCCGCTGCACAAGGTTTTCAGGCATGCCGTTTTGTTGCTTTTGGCTTACAGGATTGTGTAACCGGTATTAATGATGATGATTATGTCATTGCATTGACTGGTACTAATCAGCTCTGTGCCAAAATTTTACCTTTTTCTGATAGACCCCTTAATTT GCGAGGTTGGCTCATTTTTT-3’ (GenBank accession number: AY567487.2)
5) The sequence of standard sample for the ORF1a gene of TGEV (Integrated DNA technologies, USA): 5’-AAAGTGAGTGTAGCGTGGCTATATCTCTTCTTTTACTTTAACTAGCCTTGTGCTAGATTTTGTCTTCGGACACCAACTCGAACTAAACGAAATATTTGTCTTTCTATGAAATCATAGAGGACAAGCGTTGATTATTTCCATTCAGTTTGGCAATCACTCCTTGGAACGGGGTTGAGCGAACGGTGCAGTAGGGTTCCGTCCCTATTTCGTAAGTCGCCTAGTAGTAGCGAGTGCGGTTCCGCCCGTACAACGTTGGGTAGACCGGGTTCCGTCCTGTGATCTCCCTCGCCGGCCGCCAGGAGAATGAGTTCCAAACAATTCAAGATCCTTGTTAATGAGGACTATCAAGTCAACGTGCCTAGTCTTCCTATTCGTGACGTGTTACAGGAAATTAAGTACTGCTACCGTAATGGATTTGAGGGCTATGTTTTCGTACCAGAATACTGTCGTGACCTAGTTGATTGCGATCGTAAGGAT CACTACGTCATTGGTGTTCTTGG-3’ (GenBank accession number: KX900394.1)
6) The sequence of standard sample for the L5 gene of AdV (Integrated DNA technologies, USA): 5’-CAAAATGTAACCACTGTTACTCAGCCACTTAAAAAAACAAAGTCAAACATAAGTTTGGACACCTCCGCACCACTTACAATTACCTCAGGCGCCCTAACAGTGGCAACCACCGCTCCTCTGATAGTTACTAGCGGCGCTCTTAGCGTACAGTCACAAGCCCCACTGACCGTGCAAGACTCCAAACTAAGCATTGCTACTAAAGGGCCCATTACAGTGTCAGATGGAAAGCTAGCCCTGCAAACATCAGCCCCCCTCTCTGGCAGTGACAGCGACACCCTTACTGTAACTGCATCACCCCCGCTAACTACTGCCACGGGTAGCTTGGGCATTAACATGGAAGATCCTATTTATGTAAATAAT-3’ (GenBank accession number: J01917.1)

### **Text S3. Propagation of testing viruses**

We selected five viruses for this study: Tulane virus (TV, *caliciviridae*, a viral surrogate for human norovirus (Yu et al., 2013)), rotavirus (RV, *Reoviridae*), adenovirus (AdV, *Adenoviridae*), human coronavirus (NL63, *coronaviridae*), and porcine coronavirus (TGEV, *coronaviridae*). TV was propagated in MA104 cells (CRL-2378.1, ATCC, USA) supplemented by complete culture medium, which included 1X minimum essential medium (MEM; Thermo Fisher Scientific, USA), 2% fetal bovine serum (FBS; Thermo Fisher Scientific, USA), 1X antibiotic-antimycotic (Thermo Fisher Scientific, USA), 17 mM of NaHCO_3_, 10 mM of HEPES, and 1 mM of sodium pyruvate (Thermo Fisher Scientific, USA). After 2 days incubation at 37℃ with 5% CO_2_, TV was harvested by three cycles of freeze and thaw. TV was purified by centrifugation at 2000 rpm (556 g) for 10 min (Sorvall Legend RT Plus, Thermo Fisher Scientific, MA, USA), followed by filtration through a 0.22 μm filter (Millipore Sigma, MA, USA). RV was first activated with trypsin at a final concentration of 10 μL/mL for 1 hour in a 37℃ water bath. Then, the viruses were inoculated in MA104 cells with the culture medium without any serum (i.g., 0% FBS) and trypsin was also added at a final concentration of 2 μL/mL. Cytopathic effect appeared after 3-day incubation at 37℃ with 5% CO_2_, after which the viruses were harvested and purified in the same way described for TV.  AdV was propagated in A549 (CCL-185, ATCC) and grown in Ham F-12 media with 10% fetal bovine serum (FBS; Thermo Fisher Scientific, MA, USA) and 1X antibiotic-antimycotic (Thermo Fisher Scientific, MA, USA). They were incubated at 37℃ with 5% CO_2_ for 5 days, and then the viruses were harvested and purified in the same method for TV and RV. We inoculated NL63 in MK2 cells (CCL-7, ATCC) and used the same complete culture medium for TV. We grow the viruses at 32℃ with 5% CO_2_ for 6 days (note that the MK2 cells were grown at 32℃ with 5% CO_2_ until they became 90-100% confluent). After that, the supernatant of the virus inoculum was taken without any freezing and thawing cycle. Then, the viruses were purified by centrifugation and filtration, which is the same for TV.  TGEV was propagated in ST cells (CRL-1746, ATCC). The same MEM-based culture medium for TV and MK2 was used to grow TGEV. The viruses were incubated at 37℃ with 5% CO_2_ for 2 days. Virus harvest and purification followed the same method for TV.

### **Text S4. Virus titer measurement**

Plaque assays were used to determine titers of TV, RV, AdV, and TGEV. TCID50 assays were used to quantify NL63 titers. MA104 (ATCC, CRL-2378.1) was used as a host cell line for TV and RV propagation. MA104 was supplemented by the culture medium containing 1X minimum essential medium (MEM; Thermo Fisher Scientific, MA, USA), 10% fetal bovine serum (FBS; Thermo Fisher Scientific, MA, USA), 1X antibiotic-antimycotic (Thermo Fisher Scientific, MA, USA), 17 mM of NaHCO_3_, 10 mM of HEPES, and 1 mM of sodium pyruvate. For plaque assays using TV, 800 μL TV-containing solution was added to one well of a >90% confluent MA104 monolayer in 6-well plates (USA Scientific, USA). Viruses were incubated with the monolayer for 1 hour at 37℃ and 5% CO_2_. When performing dilutions, the virus solution was diluted 10-fold in MEM-based culture medium lacking 10% FBS. After one hour, supernatants were aspirated, and 2 mL of an overlay solution [1X minimum essential medium (MEM; Thermo Fisher Scientific, MA, USA), 1% agarose, 7.5% sodium bicarbonate, 15 mM HEPES, and 1X antibiotic-antimycotic (Thermo Fisher Scientific, MA, USA)], were added to the cellular monolayers. The overlay solution was solidified at 4℃ for 20 minutes. The solidified overlay was removed after 2-day incubation at 37℃ with 5% CO_2_. Then, 2 mL of a 10% formaldehyde solution was added to a well, and incubated at room temperature for about one hour. The solution was removed and plaques were visualized. Plaque assay to quantify TGEV used a protocol identical to above except that the ST cell line (ACC, CRL-1746) was used instead of the MA104 cell line. Note that ST cells were grown in the same culture medium as that for MA104 cells. Plaque assays to quantify RV was similar to that for TV. However, because trypsinization of RV is required for RV to attach to and infect MA104 cells, the overlay solution included an additional component of trypsin at 1 µg/mL, and the incubation time was 4 days instead of 2 days before the formaldehyde solution was added. Plaque assays to quantify AdV used 90% confluent A549 (ATCC; CCL-185) cellular monolayers (Oh et al., 2020). A549 cells were cultured in Ham F-12 media containing 10% FBS (Thermo Fisher Scientific, MA, USA), and 1X antibiotic-antimycotic (Thermo Fisher Scientific, MA, USA). In this case, 400 μL of each AdV dilution was inoculated onto cellular monolayers in six-well plates. Serial dilutions of AdV occurred in Ham's F12-K media lacking 10% FBS. After an adsorption phase, virus inoculum was removed and cellular monolayers were incubated with 2 mL of overlay solution containing 2X minimum essential medium (Thermo Fisher Scientific, MA, USA), antibiotic-antimycotic solution, 0.05 mL of 15 mM HEPES, 0.03 mL of 7.5% sodium bicarbonate, 0.5 mL of 1% agarose solution, and 0.1 mL of FBS. Plates were incubated at 4°C for about 20 min to solidify the agar overlay. Next, plates were incubated at 37°C with 5% CO_2_ for 5 days. The cell attachment and dyeing processes were the same as described for TV.

MK2 cells (ATCC, CCL-7) were used to determine NL63 titers. In this case, due to the lack of clear cytopathic effect (CPE), we developed a new TCID50 assay that integrates cell culture and RT-qPCR to quantify NL63. MK2 cellular monolayers were plated in 96-well plates. 20 µL NL63 samples were diluted in 180 µL of the MEM solution, and inoculated to each well. The 5 µL of initial virus solution on the well was taken from each sample and stored at -80 C, which was used as controls. The inoculum was incubated at 32℃ with 5% of CO_2_ for 6 days, which was determined based on the NL63 growth curve (**SI Fig. 2**). The gap between the cover and the plate was sealed with tape to prevent medium evaporation. After the incubation, 5 µL of medium was collected from each sample and mixed with 45 µL of 100-fold proteinase K solution. The viral genomes were extracted by the heat denaturation method. The control samples were also subjected to the same heat denaturation process. The Ct values were obtained from each sample and controls by RT-qPCR. We assumed if there are infectious NL63 in the inoculum, Ct value will decrease 6 days after the infection due to new prozenies. We conducted the TCID50 assays with 10-fold serial dilutions of NL63 (36 samples in total) and found the samples are divided into two groups at ΔC_t_ of 2 (**Fig. S2**). We believe the increase in Ct after 6-day incubation is because infectious viruses successfully replicate their genomes while the similar or decreased Ct value is because viruses cannot produce prozenies. We assumed our TCID50 assay reads virus sample positive (or infectious) when ΔC_t_ is larger than 2. Thus, we decided ΔC_t_ of 2 as an indicator for positive NL63 infection (**Fig. S3**). We tested four molecular replicates of 10-fold serial dilutions, each of which has two technical replicates, for one sample. TCID50 was calculated by Reed–Muench Method with the number of positive and negative samples (Lei et al., 2021).

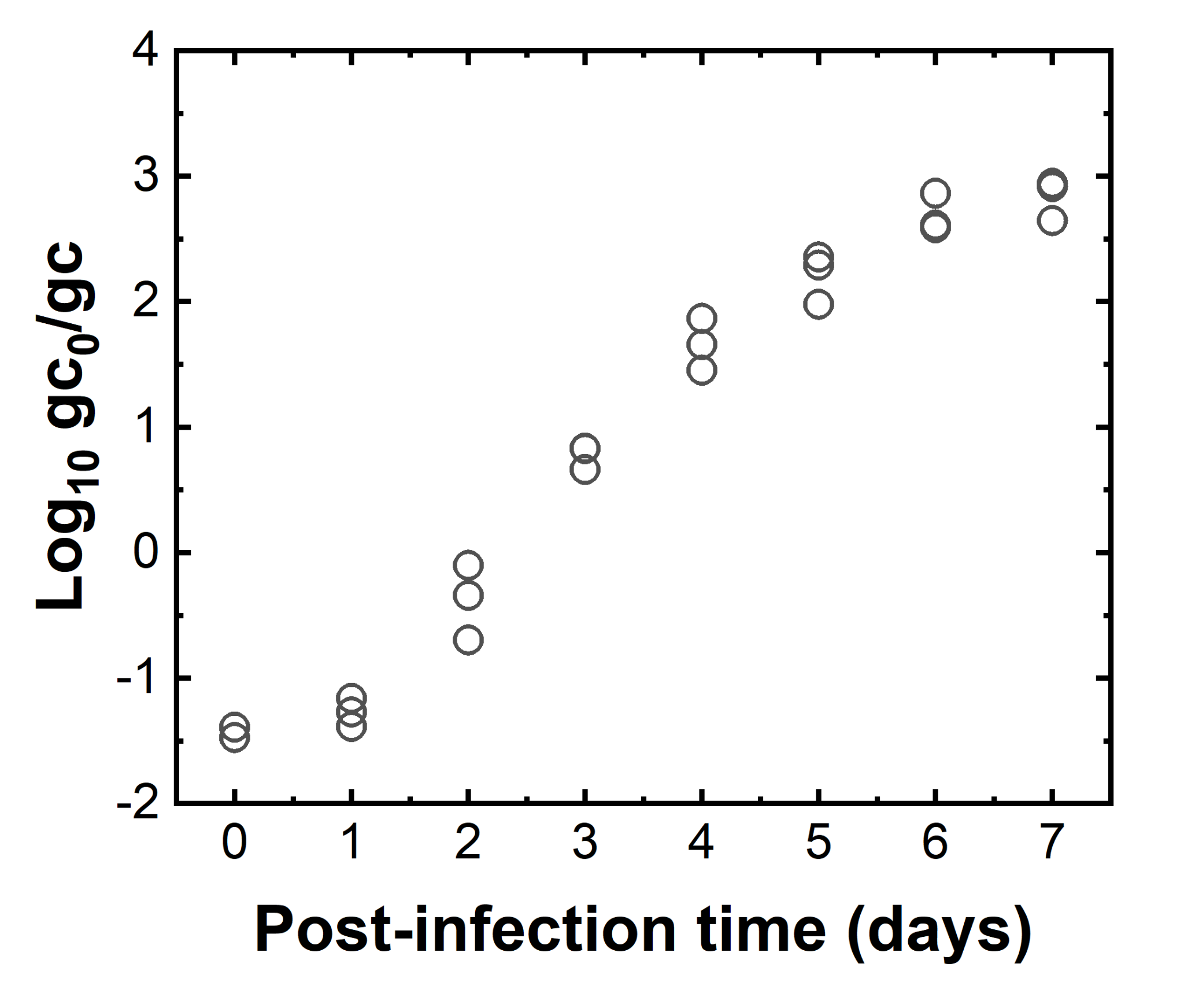

**Fig. S2.** A growth curve of NL63 on MK2 cells. One-hundred microliter of NL63 was inoculated to a monolayer of MK2 cells on 6-well plates with a multiplicity of infection (MOI) of 0.1. The inoculated viruses were incubated at 32℃ with 5% CO_2_ for 2 hours to promote virus attachment to the host cells. The inoculum was removed and the cells were rinsed three times by PBS to remove unbound virus particles. The cells were supplemented by 800 µL of culture media and incubated at 32°C with 5% CO_2_ for 7 days. The number of NL63 genomes in supernatant (Log_10_ gc) was quantified everyday and normalized to that in the initial inoculum (Log_10_ gc_0_).

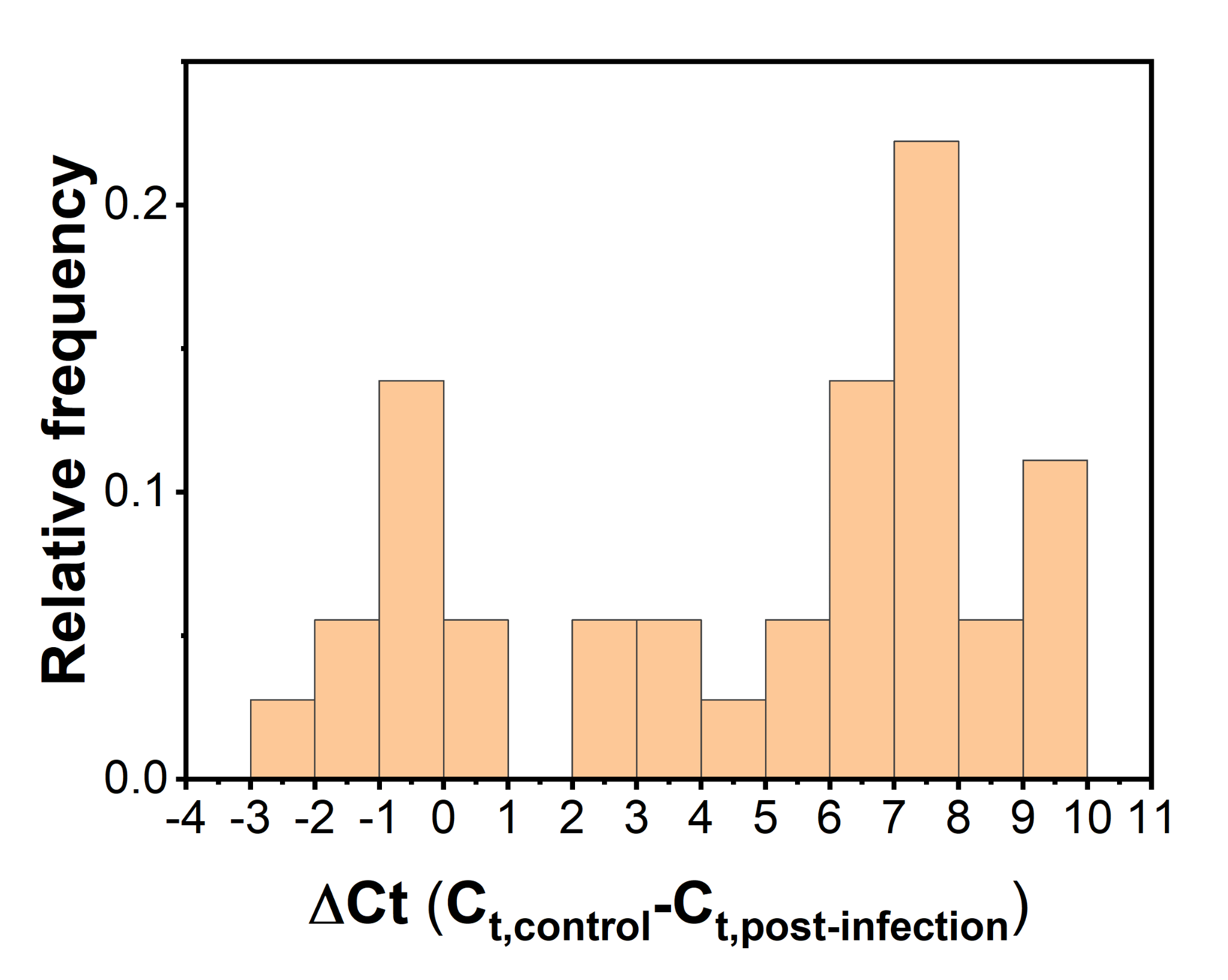

**Fig. S3.** A histogram of TCID50 assay results with 10-fold serial dilutions of NL63 (1, 10, and 100-fold dilution). MK2 cells were prepared on 96-well plates with 180 µL of culture medium supplemented. NL63 (20 µL) was inoculated to the MK2 cells and incubated at 32°C with 5% CO_2_ for 6 days. ΔC_t_ was calculated by subtracting C_t_ values of NL63 after the incubation period (C_t,post-infection_) from C_t_ values in the virus inoculum (C_t,control_). In total, 36 samples were analyzed in the histogram with 13 bins. The samples are divided into two groups at ΔC_t_ of 2, and thus we used ΔC_t_ of 2 for criterion to determine positives for infectious NL63.

**Table S3.** Summary of sewage samples collected across the Champaign County, IL

| Sample name | Location | Date | Note | Data |
| --- | --- | --- | --- | --- |
| WW | Urbana | Feb-3-2021 | Influent wastewater | Fig. 1, Fig. 2, Fig. 4, Table 2 |
| PM | Champaign | Jan-25-2021 | Residential sewage | Fig. 7 |
|  |  | Mar-22-2021 |  | Fig. 7 |
|  |  | Apr-19-2021 |  | Fig. 7 |
|  |  | Apr-26-2021 |  | Fig. 7 |
|  |  | May-3-2021 |  | Fig. 7 |
|  |  | May-10-2021 |  | Fig. 7 |
|  |  | Jun-13-2021 |  | Table 3 |
|  |  | Jun-21-2021 |  | Table 3 |
| CT | Champaign | Mar-29-2021 | Residential sewage | Fig. 7 |
|  |  | May-3-2021 |  | Fig. 7 |
|  |  | May-10-2021 |  | Fig. 7 |
|  |  | Jun-21-2021 |  | Fig. 5, Fig. 6 |
| RW | Champaign | May-3-2021 | Residential sewage | Fig. 7 |
|  |  | Jun-1-2021 |  | Table 3 |
| WS | Champaign | Jun-1-2021 | Residential sewage | Table 3 |
| GF | Champaign | Jun-1-2021 | Residential sewage | Fig. 5, Fig. 6 |
| PP | Rantoul | Jan-27-2021 | Industrial wastewater | Fig. 7 |
|  |  | Feb-10-2021 |  | Fig. 7 |
|  |  | Mar-3-2021 |  | Fig. 7 |
|  |  | Apr-14-2021 |  | Fig. 7 |
|  |  | Apr-21-2021 |  | Fig. 7 |
|  |  | May-12-2021 |  | Fig. 7 |
|  |  | Jun-30-2021 |  | Fig. 5, Fig. 6 |
| ST | Rantoul | Apr-28-2021 | Residential sewage | Fig. 7 |
|  |  | Jul-7-2021 |  | Fig. 5, Fig. 6 |
|  |  | Jun-2-2021 |  | Table 3 |
|  |  | Jul-7-2021 |  | Table 3 |
| TP | Rantoul | Apr-21-2021 | Residential sewage | Fig. 7 |
| MT | Rantoul | Jun-2-2021 | Residential sewage | Fig. 5, Fig. 6 |
|  |  | Jun-16-2021 |  | Fig. 5, Fig. 6 |
|  |  | May-12-2021 |  | Fig. 7 |
|  |  | Jun-9-2021 |  | Table 3 |
| LG | Rantoul | May-19-2021 | Lagoon | Fig. 7 |
| HA | In laboratory | May-19-2021 | Humic acid in water | Fig. 7 |

**Table S4.** The limit of quantification and multiplication factor of the PGM-MBs method for different volumes of initial solutions

| **Viral species** | **Parameter** | **Measured** | **Calculated** | | |
| --- | --- | --- | --- | --- | --- |
|  |  | **10** | **50** | **200** | **1000** |
| **TV** | LOQs | 5.10 | 4.40 | 3.79 | 3.10 |
|  | Multiplication factor | 15.2±9.2 | 76 | 304 | 1520 |
| **RV** | LOQs | 5.57 | 4.88 | 4.27 | 3.57 |
|  | Multiplication factor | 12.3±4.7 | 61.5 | 246 | 1230 |
| **AdV** | LOQs | 4.63 | 3.93 | 3.33 | 2.63 |
|  | Multiplication factor | 64.0±16.6 | 320 | 1280 | 6400 |
| **NL63** | LOQs | 5.62 | 4.92 | 4.32 | 3.62 |
|  | Multiplication factor | 10.3±2.3 | 51.5 | 206 | 1030 |
| **TGEV** | LOQs | 6.08 | 5.38 | 4.78 | 4.08 |
|  | Multiplication factor | 1.3±0.5 | 6.5 | 26 | 130 |

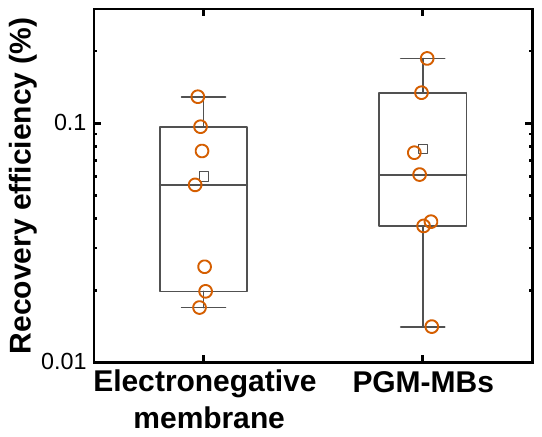

**Fig. S4**. Recovery efficiencies for NL63 by the electronegative membrane filtration and the PGM-MBs methods. Each box plot consists of 7 different samples that were collected from either different locations or dates. Each group of data and the difference between the paired samples are normally distributed. The paired sample t-test presented there is no significant difference in NL63 recovery efficiency by the electronegative membrane filtration and the PGM-MBs methods (p>0.05).

**Table S5.** Performances of conventional virus concentration methods

| Reference | Extraction method | Testing viruses | CF | RE | MF |
| --- | --- | --- | --- | --- | --- |
| LaTuner et al. (2021) | Direct extraction | BCoV | 0.3333 | 0.04 | 0.1 |
|  | Electronegative membrane | BCoV | 0.0040 | 0.01 | 2.4 |
|  | Electronegative membrane | BCoV | 0.0333 | 0.01 | 0.2 |
|  | PEG | BCoV | 0.0033 | 0.00 | 0.2 |
|  | Ultrafiltration | BCoV | 0.0100 | 0.00 | 0.4 |
| Ye et al., (2016) | PEG | MS2 | 0.0043 | 0.42 | 98.0 |
|  | PEG | MHV | 0.0043 | 0.04 | 9.3 |
|  | Ultracentrifuge | MS2 | 0.0043 | 0.04 | 9.3 |
|  | Ultracentrifuge | MHV | 0.0043 | 0.05 | 11.7 |
|  | Ultrafiltration | MS2 | 0.0043 | 0.85 | 198.3 |
|  | Ultrafiltration | MHV | 0.0043 | 0.50 | 116.7 |
|  | Ultrafiltration | T3 | 0.0043 | 0.25 | 58.3 |
|  | Ultrafiltration | phi6 | 0.0043 | 0.17 | 39.7 |
| Philo et al., (2021) | BMFS | OC43 | 0.0008 | 0.29 | 357.7 |
|  | BMFS-VERTREL | OC43 | 0.0004 | 0.03 | 74.0 |
|  | PEG | OC43 | 0.0090 | 0.03 | 3.1 |
|  | Skimmed Milk | OC43 | 0.0043 | 0.05 | 10.7 |
|  | Skimmed Milk-VERTREL | OC43 | 0.0034 | 0.02 | 5.0 |
|  | Ultrafiltration | OC43 | 0.0010 | 0.01 | 8.2 |
| Ahmed et al., (2020) | Electronegative membrane | MHV | 0.0029 | 0.27 | 92.7 |
|  | Electronegative membrane | MHV | 0.0029 | 0.61 | 210.1 |
|  | Electronegative membrane | MHV | 0.0029 | 0.66 | 228.1 |
|  | Ultrafiltration | MHV | 0.0029 | 0.56 | 194.4 |
|  | Ultrafiltration | MHV | 0.0029 | 0.28 | 97.2 |
|  | PEG | MHV | 0.0058 | 0.44 | 76.4 |
|  | Ultracentrifugation | MHV | 0.0029 | 0.34 | 116.3 |
| Randazzo et al., (2020) | adsorption-precipitation | MgV | 0.0083 | 0.12 | 14.8 |
|  |  |  | 0.0100 | 0.25 | 25.4 |
|  |  |  | 0.0183 | 0.21 | 11.7 |
|  |  |  | 0.0050 | 0.08 | 16.6 |
|  |  |  | 0.0117 | 0.04 | 3.8 |
|  |  |  | 0.0117 | 0.47 | 40.7 |
|  |  |  | 0.0117 | 0.17 | 14.1 |
|  |  |  | 0.0117 | 0.33 | 28.3 |
|  |  |  | 0.0183 | 0.62 | 33.6 |
|  |  |  | 0.0100 | 0.73 | 73.1 |
|  |  |  | 0.0083 | 0.05 | 5.4 |
|  |  |  | 0.0117 | 0.17 | 14.7 |
|  |  |  | 0.0233 | 0.17 | 7.4 |
|  |  |  | 0.0100 | 0.07 | 7.5 |
|  |  |  | 0.0083 | 0.58 | 69.3 |
|  |  |  | 0.0083 | 0.26 | 31.1 |
|  |  |  | 0.0100 | 0.10 | 10.4 |
|  |  |  | 0.0117 | 0.20 | 17.1 |
|  |  |  | 0.0100 | 0.04 | 3.5 |
|  |  |  | 0.0100 | 0.07 | 7.0 |
|  |  |  | 0.0100 | 0.01 | 1.4 |
|  |  |  | 0.0067 | 0.03 | 4.0 |
|  |  |  | 0.0100 | 0.03 | 3.0 |
|  |  |  | 0.0100 | 0.16 | 16.0 |
|  |  |  | 0.0125 | 0.03 | 2.3 |
|  |  |  | 0.0120 | 0.08 | 6.6 |
|  |  |  | 0.0100 | 0.02 | 1.8 |
|  |  |  | 0.0100 | 0.02 | 2.0 |
|  |  |  | 0.0117 | 0.01 | 1.0 |
|  |  |  | 0.0133 | 0.08 | 5.8 |
|  |  |  | 0.0125 | 0.08 | 6.2 |
|  |  |  | 0.0133 | 0.01 | 0.8 |
|  |  |  | 0.0117 | 0.03 | 2.8 |
|  |  |  | 0.0117 | 0.03 | 2.6 |
|  |  |  | 0.0117 | 0.01 | 1.2 |
|  |  |  | 0.0100 | 0.02 | 2.3 |
|  |  |  | 0.0125 | 0.11 | 9.1 |
|  |  |  | 0.0100 | 0.02 | 1.9 |
|  |  |  | 0.0117 | 0.42 | 35.7 |
|  |  |  | 0.0117 | 0.42 | 35.7 |
|  |  |  | 0.0125 | 0.01 | 0.9 |
|  |  |  | 0.0117 | 0.01 | 1.1 |
|  |  |  | 0.0073 | 0.05 | 6.3 |
|  |  |  | 0.0108 | 0.02 | 1.8 |
|  |  |  | 0.0117 | 0.12 | 10.1 |
|  |  |  | 0.0117 | 0.20 | 17.5 |
|  |  |  | 0.0117 | 0.01 | 1.0 |
|  |  |  | 0.0093 | 0.06 | 6.3 |
|  |  |  | 0.0083 | 0.07 | 7.9 |
|  |  |  | 0.0100 | 0.03 | 3.2 |
|  |  |  | 0.0083 | 0.03 | 4.1 |
|  |  |  | 0.0083 | 0.02 | 2.0 |
|  |  |  | 0.0090 | 0.01 | 1.1 |
|  |  |  | 0.0105 | 0.08 | 7.9 |
|  |  |  | 0.0117 | 0.11 | 9.4 |
|  |  |  | 0.0075 | 0.07 | 8.8 |
|  |  |  | 0.0100 | 0.08 | 7.6 |
|  |  |  | 0.0092 | 0.02 | 1.8 |
|  |  |  | 0.0092 | 0.05 | 5.3 |
|  |  |  | 0.0100 | 0.15 | 15.1 |
|  |  |  | 0.0100 | 0.05 | 5.3 |
|  |  |  | 0.0133 | 0.05 | 3.5 |
|  |  |  | 0.0092 | 0.03 | 3.1 |
|  |  |  | 0.0100 | 0.05 | 4.6 |
|  |  |  | 0.0083 | 0.03 | 3.8 |
|  |  |  | 0.0117 | 0.04 | 3.4 |
|  |  |  | 0.0150 | 0.06 | 3.7 |
|  |  |  | 0.0117 | 0.04 | 3.5 |
|  |  |  | 0.0075 | 0.01 | 1.3 |
|  |  |  | 0.0092 | 0.02 | 2.6 |
|  |  |  | 0.0083 | 0.03 | 3.4 |
|  |  |  | 0.0067 | 0.12 | 18.6 |
|  |  |  | 0.0100 | 0.13 | 13.3 |

**Table S6.** Consumables’ prices for production of the PGM-MBs (accessed on Aug 10, 2021)

| Process | Item | Vendor  (Catalog number) | Price (USD) | Presentation | Price per 10 mL wastewater analysis |
| --- | --- | --- | --- | --- | --- |
| PGM-MBs synthesis | carboxyl derivatized beads | Thermo Fisher Scientific (21353) | 202 | 5 mL | 0.404 |
|  | porcine gastric mucin | Millipore Sigma (M1778) | 399 | 100 g | 0.004 |
|  | MES buffered Saline | Thermo Fisher Scientific (28390) | 152 | 10 packs | 0.003 |
|  | EDC | Thermo Fisher Scientific (22981) | 378 | 25 g | 0.002 |
| Genome extraction | Proteinase K | New England Biolabs (P8107S) | 85 | 2 mL | 0.043 |
| Total | | - | - | - | 0.456 |

**Reference**

Araud, E., Shisler, J.L., Nguyen, T.H., 2018. Inactivation Mechanisms of Human and Animal Rotaviruses by Solar UVA and Visible Light. Environ. Sci. Technol. 52, 5682–5690. https://doi.org/10.1021/acs.est.7b06562

Fuzawa, M., Araud, E., Li, J., Shisler, J.L., Nguyen, T.H., 2019. Free Chlorine Disinfection Mechanisms of Rotaviruses and Human Norovirus Surrogate Tulane Virus Attached to Fresh Produce Surfaces. Environ. Sci. Technol. 53, 11999–12006. https://doi.org/10.1021/acs.est.9b03461

Lei, C., Yang, J., Hu, J., Sun, X., 2021. On the Calculation of TCID50 for Quantitation of Virus Infectivity. Virol. Sin. 36, 141–144. https://doi.org/10.1007/S12250-020-00230-5/TABLES/2

Oh, C., Sun, P.P., Araud, E., Nguyen, T.H., 2020. Mechanism and efficacy of virus inactivation by a microplasma UV lamp generating monochromatic UV irradiation at 222 nm. Water Res. 186. https://doi.org/10.1016/j.watres.2020.116386

Yu, G., Zhang, D., Guo, F., Tan, M., Jiang, X., Jiang, W., 2013. Cryo-EM Structure of a Novel Calicivirus, Tulane Virus. PLoS One 8, 59817. https://doi.org/10.1371/journal.pone.0059817
